# Beyond Intensity: Cross-Dataset Consistency of Temporal Facial Action-Unit Dynamics as Transferable Markers of Depression

**DOI:** 10.64898/2026.07.12.26357331

**Authors:** Inyong Jeong, MyeongGyun Jang, Ju-wan Kim, Hayeon Kim, Soohyun Park, Dae-Kwang Kim, Jin-Hyun Park, Yeongmin Kim, Jae-Min Kim, Hwamin Lee, Min Jhon

**Author notes:** Corresponding authors: Jae-Min Kim; Hwamin Lee, and Min Jhon. Inyong Jeong, MyeongGyun Jang, and Ju-wan Kim contributed equally to this work.

## Abstract

Facial behavior is a widely studied objective signal for depressive-symptom analysis, yet most systems are trained and evaluated within a single dataset, leaving it unclear whether the learned representations reflect depression or dataset-specific artifacts. Prior work has also relied on aggregate intensity statistics and small samples. We reframe the problem by asking which facial action unit (AU) features transfer across datasets, rather than which maximize within-dataset discrimination. We used time-resolved AU dynamics from a Korean cohort of 2,608 participants, including 265 with PHQ-9-defined depressive symptoms, recorded under happy and unhappy emotion-elicitation conditions. From AU time-series extracted with OpenFace, we computed 568 features spanning intensity, temporal, dynamic, peak-structure, and Duchenne (genuine smile) co-activation patterns. We then tested their directional transferability on the US DAIC-WOZ dataset, which differs in race, language, task, recording length, and setting. Within the source cohort, peak-interval features gave the strongest signal (AU26 peak-interval, Cohen’s d = −0.66), yet reversed sign externally, whereas slower temporal features and AU06 (cheek raiser) preserved their direction despite modest effect sizes. Excluding peak features raised directional agreement from 56-64% to 82-90%. Within-dataset discriminative strength therefore does not predict cross-dataset transferability, supporting directional consistency as a practical criterion for selecting transferable affective-model inputs. Smile analysis further showed that depressive symptoms were marked less by reduced smiling than by eye-mouth decoupling (d = −0.41), which survived covariate adjustment and matched the voluntary–emotional facial-pathway distinction. Being low-dimensional and non-identifying, AU time series may support privacy-conscious multisite depression research without raw video.

## I. INTRODUCTION

### A. Background

Depression affects approximately 332 million people worldwide [1]. Despite this burden, many affected individuals never receive timely care, as stigma, limited mental-health literacy, and barriers to reaching specialists leave a large proportion of cases unidentified [2]. Identifying depressive symptoms from objective, automatically measurable data has therefore emerged as a potential way to broaden early identification and reach people who might otherwise go undetected. Machine-learning models have been applied to detect depression-related signals from such data, including speech, physiological signals, and facial behavior [3–5], yielding objective markers that can complement conventional assessment.

Facial expression is a strong candidate for such a marker. Cohn et al. compared automatically measured facial actions with clinical diagnosis in patients treated for depression and reached 88% accuracy with manual FACS coding and 79% with automatic appearance modeling, comparable to vocal prosody [6]. Girard et al. followed depressed patients across treatment and found that facial behavior tracked symptom severity over time, with manual and automatic coding showing close agreement [7]. These findings indicate that facial behavior carries an objective signal of depression that automated tools can read at scale. Quantifying that signal requires a way to describe facial movement in consistent units. The Facial Action Coding System (FACS) of Ekman and Friesen meets this need by encoding the movement of individual facial muscles as action units (AUs), providing a quantitative description of facial expression [8, 9]. Automatic tools such as OpenFace extract AU intensity frame by frame from video, making facial analysis feasible in large samples [10].

Studies using FACS have reported a recurring pattern in how specific AUs change with depression. Higher symptom severity is associated with fewer affiliative expressions, particularly reduced AU12 (lip corner puller), and with more non-affiliative expressions such as AU14 (dimpler), reflecting a loss of positive expressivity that has been interpreted as social withdrawal [6, 7]. These findings, however, come mainly from single datasets and rely on aggregate measures such as mean AU intensity, and for some units the reported associations are not stable. AU06 (cheek raiser), the eye component of a genuine smile, has been associated with the non-depressed state in some cohorts [11] but reported as non-significant or higher in depressed participants in others [12], with reduced activation of positively valenced units also observed in related psychiatric populations [13]. This instability may reflect, in part, heterogeneity among samples that share the same total symptom score [14] and, in part, the limitations of comparing single mean values across small datasets. Such inconsistency motivates testing whether a larger sample and time-resolved features can establish the direction of AU effects more reliably, rather than leaving it unclear whether a given pattern reflects depression itself or the particular dataset in which it was measured.

The quality of a smile offers an additional cue. A Duchenne smile, the genuine form, co-activates AU06 and AU12, whereas a non-Duchenne smile activates AU12 alone [15]. AU12 is under voluntary control and can be produced on demand, while AU06 accompanies genuine positive affect involuntarily and is difficult to suppress or fake [16]. The clinical observation that Duchenne smiles decline in depression is long-standing [17], but no study has classified these two smile types frame by frame in a large sample or tracked how they change over time and across emotional contexts.

### B. Open Challenges and Our Approach

Research linking AUs to depression faces three limitations. First, samples are small. DAIC-WOZ, the standard dataset for facial depression research, contains 189 interviews [18], and facial studies in Korean cohorts have involved only tens of participants [19]. Models fitted to a small single dataset reach high accuracy under controlled validation but tend to overfit, and overfitting remains a recognized concern even within standardized benchmark settings [20]. Models trained on standard datasets have even been shown to capture general psychological distress rather than features specific to depression [21], which makes external validation of single-dataset results necessary. Second, features are dominated by aggregate measures such as the mean and standard deviation. How facial behavior changes across a session, through segment-wise intensity, burst frequency, autocorrelation, or peak intervals, has rarely been examined. Parikh et al. attempted temporal analysis of AUs in DAIC-WOZ but stopped at the mean and standard deviation [12]. Fisher et al. showed that within-session and between-session changes in facial expression run in different directions, underscoring the value of temporal resolution [22]. Third, few studies have tested whether a marker found in one dataset holds in the same direction across a different race, language, task, and recording setting. External validation, when done, has usually stayed within the same US dataset family (DAIC-WOZ to E-DAIC) [23]. Because norms of emotional expression differ across cultures [24], confirming whether an AU-depression association found in a Korean cohort reproduces in US data is essential for judging generalizability. A few recent speech- and text-based studies have begun to test depression detection across languages, reporting that models transfer unevenly across English, Chinese, Korean, and German data [25, 26], but such work has focused on whole-model classification performance rather than on which specific markers preserve their direction, and it has not addressed facial behavior.

These three limitations share a common bottleneck that constrains the scaling of facial psychiatric AI. Identifiable facial video is hard to share across institutions and countries because of privacy risk, which impedes multi-site training and external validation [27]. AU intensity time-series are low-dimensional signals extracted from video; they carry lower re-identification risk and are far smaller than raw video, so they are easier to share. Identifying which AU features are consistently associated with depression across datasets is therefore tied directly to the problem of finding transferable inputs that do not require sharing the video itself. Yet which AU features transfer across datasets and which are artifacts of a particular dataset has not been systematically separated.

Using a large Korean cohort, we recorded facial expression under happy and unhappy emotion-elicitation conditions and used time-resolved AU features rather than aggregate statistics to address three questions. First, we test whether AU differences between PHQ-9 screen-positive and screen-negative participants reproduce the findings of earlier small studies. Second, we evaluate whether these differences hold in the same direction in the US DAIC-WOZ data, which differ in race, language, task, and recording setting, and which feature types transfer across datasets and which do not. Third, we analyze how the composition and temporal course of Duchenne-type versus AU12-only smile patterns differ in depressive symptoms, and we ask whether eye-mouth decoupling provides a mechanistically interpretable behavioral marker that is consistent with, but does not directly prove, the known separation of emotional and volitional pathways for facial expression.

## II. METHODOLOGY

### A. Study Cohort and Data Collection

This study was based on a single Korean cohort. Participants were recruited at Chonnam National University Hospital (CNUH) and Chonnam National University Hwasun Hospital (CNUHH) from August 2, 2021, to September 26, 2025, as part of a study designed to investigate mental disorders using a transdiagnostic approach based on common symptoms and processes. The dataset includes psychological scale scores, task-based recordings, heart rate variability (HRV), vital signs, and blood and actigraphy data. All participants provided written informed consent prior to participation. Key inclusion criteria for the parent study were being over 18 years of age and being able to wear an actigraphy device and provide digital data such as voice, facial video, and smartphone usage data.

Among the tasks in the dataset, this study focused on two free narrative tasks in which participants were asked to recall and describe in detail their happiest memory and their unhappiest memory, respectively. This task design was informed by prior work on autobiographical emotional memory recall in depression and on multimodal/behavioral analysis of psychological distress [18, 28, 29]. When needed, trained interviewers provided minimal prompts without influencing the content. All sessions were video-recorded in a quiet room under standard indoor lighting, with participants seated. Recordings were made using the rear-facing camera of a Samsung Galaxy Tab S7 tablet (Samsung Electronics, Suwon, South Korea), mounted above the monitor that the participant was facing, at an approximate distance of 60–90 cm from the participant, to capture a frontal view of the face. Videos were recorded at a resolution of 1920 × 1080 (1080p) and a frame rate of 30 fps. Only the facial video data were used in this analysis.

A total of 2,614 raw videos were collected for the happy condition and 2,616 for the unhappy condition. Some participants were excluded during video quality filtering, and the analysis included participants who had a valid video and a PHQ-9 response in both conditions. The final cohort comprised 2,608 participants, each assessed under both conditions. Depressive symptoms were assessed with the PHQ-9 [30]. The total score was computed as the sum of the nine items, and a score of 10 or above defined the PHQ-9 screen-positive group. This group included 265 participants (10.2%) and the screen-negative group included 2,343 participants. In the remainder of the article, we use “depressed group” and “non-depressed group” as shorthand for these PHQ-9-defined groups rather than as structured clinical diagnoses. Demographic characteristics of the cohort are presented in Table I. The study was approved by the CNUH and CNUHH Institutional Review Boards (approval nos. CNUH-2021-243, CNUH-2022-216, CNUHH-2021-117, and CNUHH-2022-126), and written informed consent was obtained from all participants.

**TABLE I.** BASELINE DEMOGRAPHIC AND SPEECH CHARACTERISTICS OF THE STUDY COHORT.

| Characteristic | Total (n=2,608) | Depressed (n=265) | Non-depressed (n=2,343) | p |
| --- | --- | --- | --- | --- |
| **Sociodemographic characteristics** |  |  |  |  |
| Age (year) | 63 [48–70] | 51 [30–62] | 64 [50–70] | <0.001 |
| Female, n (%) | 1,760 (67.5) | 165 (62.3) | 1,595 (68.1) | 0.065 |
| Unmarried, n (%) | 787 (30.2) | 144 (54.3) | 643 (27.4) | <0.001 |
| No religion, n (%) | 1,130 (43.3) | 140 (52.8) | 990 (42.3) | 0.001 |
| Living alone, n (%) | 453 (17.4) | 68 (25.7) | 385 (16.4) | <0.001 |
| Alcohol use (yes), n (%) | 959 (36.8) | 101 (38.1) | 858 (36.6) | 0.681 |
| Current smoker (yes), n (%) | 237 (9.1) | 56 (21.1) | 181 (7.7) | <0.001 |
| **Speech characteristics** |  |  |  |  |
| Speech length, Happy (s) | 174 [123–201] | 123 [78–176] | 177 [132–203] | <0.001 |
| Speech length, Unhappy (s) | 175 [127–204] | 150 [102–194] | 176 [130–205] | <0.001 |

Values are presented as median [interquartile range (IQR)] or n (%). Continuous variables were compared using the Mann–Whitney U test and categorical variables using the chi-square test. Unmarried includes never-married, divorced, widowed, and cohabiting participants. Speech length was calculated from the total number of recorded frames assuming a frame rate of approximately 30 frames/s.

### B. Action Unit Extraction

Frame-wise AU intensity was extracted from the videos with OpenFace 2.0 [10]. The intensities (0–5) of 17 AUs were used for analysis (Supplementary Table S1): AU01, AU02, AU04, AU05, AU06, AU07, AU09, AU10, AU12, AU14, AU15, AU17, AU20, AU23, AU25, AU26, and AU45. Frames with a confidence below 0.5 or a success value of 0 were excluded, and participants with fewer than 20 valid frames were excluded from the analysis. Because participant identifier formats differed across datasets, identifiers were standardized to link the video, the AU features, and the PHQ-9 label.

### C. Temporal AU Representation

For each of the 17 AUs, 32 features were computed from its intensity time series across all valid frames. In addition, two coordination features were extracted for each of 10 AU pairs, and four global facial-coordination features were computed. This yielded 568 features in total (17 × 32 + 20 + 4). The extracted feature definitions are summarized in Supplementary Table S2.

The first group describes the overall level of each AU, summarized by its mean and median intensity over the session. A second group describes how the AU changes across the session. To capture this at two time scales, the session was divided into thirds and the mean intensity of the early, middle (mid), and late portions was taken, and it was separately divided into five equal blocks whose means summarize the beginning (se), middle (sm), and end (sl) of the session; the finer division reflects short-term fluctuation, while the coarser division reflects slow drift. From these, several trend measures were derived, including the linear slope of intensity over time (slope) and the difference between the late and early portions (le), both expressing whether the AU strengthened or weakened as the session progressed.

A third group describes the variability of each AU, measured by its standard deviation, interquartile range, skewness, and kurtosis, capturing how widely and how asymmetrically intensity was distributed. A fourth group describes the moment-to-moment dynamics: the average size of the change between consecutive frames (roc) and its variability quantify how abruptly the AU moved, and the correlation of the signal with itself at short delays of 1, 5, and 10 frames (autocorrelation) quantifies how persistent its activity was over time.

A fifth group describes bursts of strong activity. A burst was defined as a continuous stretch in which intensity stayed above the 75th percentile, and we counted how often such bursts occurred, how long they lasted on average and at their longest, and how regularly they were spaced. A sixth group describes how often the AU switched on and off: treating intensity above the median as active, we measured how frequently it became active, what fraction of the session it was active, and how readily it transitioned between active and inactive states. A seventh group describes the rhythm of peaks: peaks in the intensity signal were detected automatically, and we recorded how many occurred (npk) and how evenly they were spaced (the mean inter-peak interval, pk_int).

The remaining features describe how AUs move together. For each of 10 AU pairs, we computed how strongly the two AUs co-varied within the same frame (correlation) and whether one tended to lead or follow the other in time. Finally, four global features summarized overall facial coordination: the average number of AUs active at the same time, and, for the Duchenne smile specifically (the joint activation of the cheek raiser AU06 and the lip corner puller AU12), how often the two co-occurred (the Duchenne rate) and how long such episodes lasted. Features based on event counts were normalized to a rate per 1,000 valid frames so that they were not confounded by differences in recording length.

### D. Smile Representation

Separately from the features above, smile quality was analyzed from the frame-wise activation of AU06 and AU12. Each AU was treated as active when it exceeded its within-session median, and each frame was classified as Duchenne (both active), non-Duchenne (AU12 active only), eye-only (AU06 active only), or neutral (both inactive) (Supplementary Fig. S1). Because activation is referenced to the within-session median, these measures reflect the qualitative composition of expression rather than its absolute amount. At the overall level, we computed the frame proportion of the four smile states, smile quality (Duchenne / (Duchenne + non-Duchenne)), the mean intensity of AU06 and AU12, and the conditional intensity of AU06 during AU12-active frames (au06_given_au12). Let *I*_06_(*t*)and *I*_12_(*t*)denote the frame-wise intensities of AU06 and AU12, respectively, and let *m*_12_denote the within-session median intensity of AU12. The conditional intensity metric was defined as

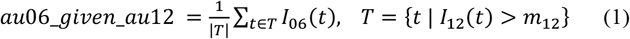

The metric was computed only when ∣T∣ > 10. It quantifies the degree to which eye activity accompanies mouth movement after controlling for differences in smile frequency. At the temporal level, each session was divided into three equal relative segments, and the Duchenne rate, non-Duchenne rate, and smile quality were computed for each segment. Because participants with depression generally spoke for shorter periods, these segments represent relative positions within a session rather than absolute time.

## III. EVALUATION

### A. Within-cohort Biomarker Analysis

For each of the 568 features, the depressed and non-depressed groups were compared using a two-sided Mann-Whitney U test. Effect size was summarized using Cohen’s d calculated with the pooled standard deviation. Multiple comparisons were corrected using both the Benjamini-Hochberg false discovery rate (FDR) procedure and Bonferroni correction. To estimate an upper bound on classification performance, features were ranked by their univariate AUROC, and the top 5, 10, 15, 20, and 30 features were entered into a logistic regression model evaluated using 5-fold stratified cross-validation. Because feature ranking was performed on the full dataset, the resulting AUROC should be interpreted as an optimistic ceiling on the available signal rather than an estimate of generalization performance. AUROC was reported as *max*(*AUC*, 1 − *AUC*) so that it reflects discriminative magnitude independent of the direction of the effect.

### B. Cross-dataset Transferability Analysis

Directional agreement was defined as the proportion of features whose Cohen’s d had the same sign across the Korean cohort and DAIC-WOZ. Let *F* denote the set of features that were FDR-significant in the Korean cohort, and let 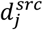 and 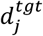 be the Cohen’s d of feature *j* in the Korean cohort and in DAIC-WOZ, respectively. The directional agreement score was defined as

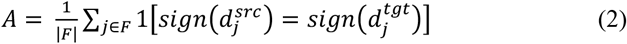

where 1[⋅]is the indicator function. Whether the observed directional agreement exceeded chance was evaluated using a binomial test. Because *F* is selected on the source cohort, *A* measures whether markers nominated in the Korean cohort hold externally, not whether the two datasets agree in general. The consistency of effect sizes between the two datasets was further quantified by correlating the Cohen’s d values across datasets, using the Pearson correlation for non-peak features and the Spearman correlation for peak features, the latter being more robust to the systematic sign reversal observed in that category. Agreement was additionally examined at both the AU level and the temporal feature-category level.

To evaluate the potential confounding effect of recording length, each feature was regressed on the log-transformed recording length, and effect sizes were recomputed using the residuals. The direction of the core AU effects was further examined across recording-length tertiles.

Demographic robustness was evaluated by examining whether the direction and statistical significance of the effects were preserved across sex, age (65-year cutoff), and recording-length (median-split) subgroups. Finally, to assess the influence of emotional context, the proportion of significant features and their effect directions were compared between the happy and unhappy conditions. Features that remained significant in both conditions with concordant effect directions were regarded as robust core biomarkers.

### C. Smile Analysis

For each of the 17 smile-related measures derived from the joint activation of AU06 and AU12, the depressed and non-depressed groups were compared using a two-sided Mann-Whitney U test and Cohen’s d, with FDR correction applied across measures. The happy and unhappy conditions were analyzed separately. All analyses were performed in Python 3.11. AU extraction used OpenFace 2.0 [10], statistical testing used SciPy 1.11, multiple-comparison correction used the multipletests function from statsmodels 0.14, and classification analyses used scikit-learn 1.3.

## IV. RESULTS

### A. Experimental Setup

Experiments consisted of two stages: within-cohort biomarker discovery using the Korean cohort (N = 2,608) and external validation using the US DAIC-WOZ dataset (N = 189).

Depression was defined as PHQ-8 ≥ 10 according to the dataset convention [18]. Because DAIC-WOZ uses the PHQ-8 rather than the PHQ-9, participants in the Korean cohort were reclassified using the PHQ-8 (sum of items 1–8, excluding suicidal ideation) with the same cutoff of 10 for the external validation analysis. The feature set remained that identified in the primary PHQ-9-based analysis, and the same features were extracted from DAIC-WOZ using an identical processing pipeline to evaluate cross-dataset transferability.

### B. Cohort Characteristics

The demographic and speech characteristics of the study cohort are summarized in Table I. The final analytic cohort comprised 2,608 participants, of whom 265 (10.2%) formed the depressed group (PHQ-9 ≥ 10) and 2,343 the non-depressed group. Compared with the non-depressed group, the depressed group was younger (median 51 vs 64 years) and more often unmarried (54.3% vs 27.4%), and had higher proportions of living alone (25.7% vs 16.4%) and current smoking (21.1% vs 7.7%) (all p < 0.001). The proportion without religious affiliation was higher among depressed participants (52.8% vs 42.3%, p = 0.001), whereas sex distribution and alcohol use did not differ significantly between groups.

In both emotion-elicitation conditions, the depressed group spoke for a shorter time (happy, 123 vs 177 s; unhappy, 150 vs 176 s; both p < 0.001). This group difference in speech length suggests that recording length could act as a potential confounder in the temporal feature analysis.

### C. Within-cohort Biomarker Discovery

Among the 568 extracted features, 66 were significant after Bonferroni correction in the happy condition and 60 in the unhappy condition (208 and 181, respectively, after FDR correction). As shown in Fig. 1, most significant features exhibited negative effect sizes, indicating lower facial activity in depressed participants. The overall pattern differed between emotional conditions. In the happy condition, significant features were dominated by peak-related measures across multiple action units, whereas in the unhappy condition they were concentrated on temporal and dynamic features derived from AU06. Across both conditions, AU06 contributed the largest number of significant features, suggesting that depression-related facial changes were primarily associated with the cheek-raiser component of facial expression.

**Fig. 1.**
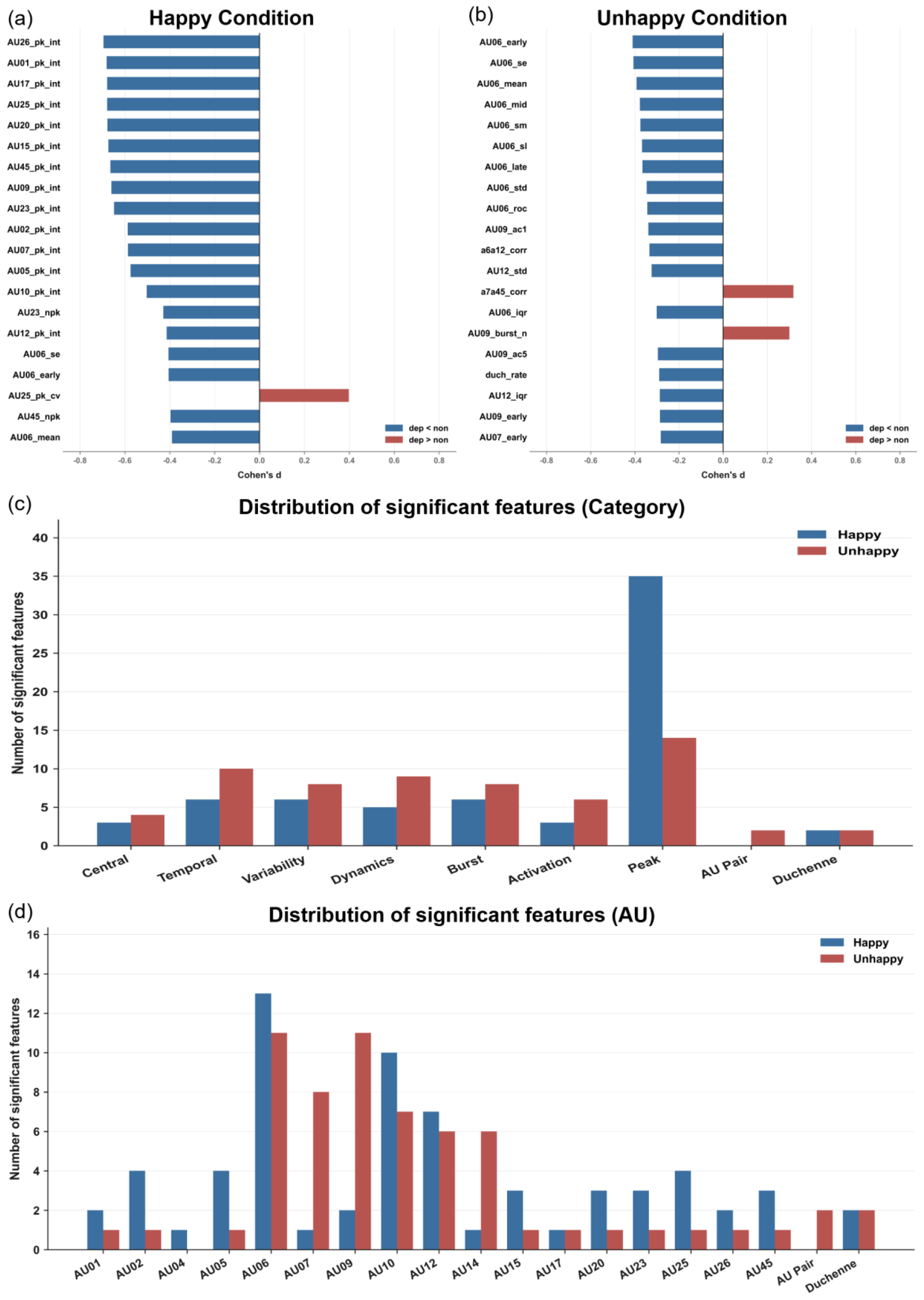
AU biomarkers distinguishing depressed and non-depressed participants. Cohen’s *d* values for Bonferroni-significant features in the happy (a) and unhappy (b) conditions. Negative values indicate lower feature values in depressed participants. Panels (c) and (d) summarize the number of significant features by feature category and by action unit (AU), respectively. Significant features in the unhappy condition were primarily derived from AU06, whereas those in the happy condition were predominantly peak-interval features

AU06 (cheek raiser) showed the strongest and most consistent association with depression. In the unhappy condition, AU06 features ranked highest in both effect size and discriminative performance (AU06_early, Cohen’s d = −0.396, AUROC = 0.615; AU06_mean, d = −0.377). The mean intensity of AU06 was lower in the depressed group under both emotional conditions (AU06_mean, d = −0.380 in happy and −0.377 in unhappy); AU12 mean intensity was in the same direction but smaller and did not survive Bonferroni correction (d = −0.232 in happy, −0.213 in unhappy).

Beyond these intensity-based measures, the strongest within-cohort signal in the happy condition came from temporal peak-interval features. The top-ranked features by both effect size and AUROC were peak-interval measures (e.g., AU26_pk_int, d = −0.662, AUROC = 0.702). These features capture the time interval between successive activation peaks of an action unit and represented the strongest discriminative signal within the Korean cohort. The multivariate upper bound on classification performance reached an AUROC of 0.72 in the happy condition and 0.65 in the unhappy condition. These values should be interpreted as an optimistic upper bound on the available depression-related facial signal rather than the expected performance of a standalone diagnostic model.

### D. Cross-dataset Transferability

We evaluated whether depression-related facial biomarkers identified in the Korean cohort preserved their effect directions in the US DAIC-WOZ dataset, which differs in race, language, interview task, recording length, and recording setting. The two datasets also differed substantially in absolute feature scale. Overall variability was greater in DAIC-WOZ, with larger standard deviations for all 17 AUs (median ratio ≈ 1.6), larger interquartile ranges for 15 of 17 AUs, and larger standard deviations of the rate of change for 16 of 17 AUs (Supplementary Table S3). In contrast, the mean intensities of the core action units AU06 and AU07 were higher in the Korean cohort. We therefore focused on whether the direction, rather than the absolute magnitude, of the depression-related effects was preserved across datasets.

Overall directional agreement reached 56% in the happy condition and 64% in the unhappy condition. However, agreement differed markedly across feature categories (Table II, Fig. 2). Temporal-segment features (happy 5/5; unhappy 9/9), central-tendency features (2/2; 3/3), and dynamic features (3/3; 6/6) showed complete directional agreement across datasets, whereas peak-related features agreed in only 10/30 (33%) in the happy condition and 0/11 (0%) in the unhappy condition. Excluding peak-related features increased directional agreement to 90% and 82%, respectively. The remaining disagreement in the unhappy condition came mainly from activation-threshold features (1/5); variability and burst features agreed in 83% of cases in both conditions.

**TABLE II.** DIRECTIONAL AGREEMENT BETWEEN THE KOREAN COHORT AND DAIC-WOZ BY FEATURE CATEGORY.

| Feature category | Happy (Agree/Total) | Agreement % | Unhappy (Agree/Total) | Agreement % |
| --- | --- | --- | --- | --- |
| Central tendency | 2/2 | 100 | 3/3 | 100 |
| Temporal segment | 5/5 | 100 | 9/9 | 100 |
| Dynamics | 3/3 | 100 | 6/6 | 100 |
| Variability | 5/6 | 83 | 5/6 | 83 |
| Burst pattern | 1/2 | 50 | 5/6 | 83 |
| Activation | 1/1 | 100 | 1/5 | 20 |
| Peak | 10/30 | 33 | 0/11 | 0 |
| Duchenne/coactivation | 1/1 | 100 | 2/2 | 100 |
| Overall | 28/50 | 56 | 32/50 | 64 |
| Overall (excluding peak) | 18/20 | 90 | 32/39 | 82 |

*Directional agreement was defined as the proportion of significant biomarkers identified in the Korean cohort whose Cohen's $d$ had the same sign in DAIC-WOZ. Categories with fewer than three significant features should be interpreted cautiously. AU-pair feature in the unhappy condition (1/2 agreement) is included in the overall total but omitted as a separate row because of the small sample size. Excluding peak-related features increased directional agreement from 56% to 90% in the happy condition and from 64% to 82% in the unhappy condition.*

**Fig. 2.**
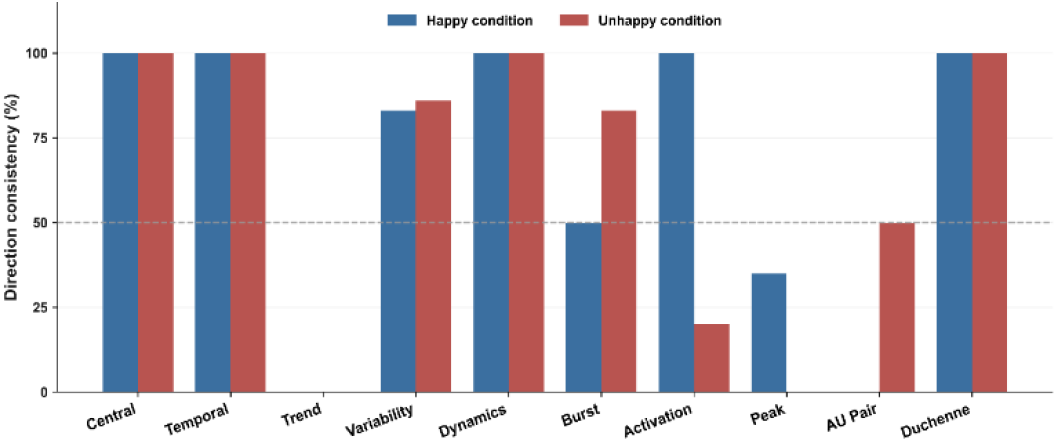
Generalization of effects to DAIC-WOZ depends on feature category. Direction agreement between the Korean cohort and DAIC-WOZ across feature categories in the happy and unhappy conditions. Temporal, central, and dynamic features show high cross-dataset consistency, whereas peak features exhibit poor agreement and systematic reversal.

The disagreement in peak-related features reflected a systematic reversal rather than random variation. In the Korean cohort, depressed participants exhibited shorter inter-peak intervals (negative Cohen’s d for pk_int), whereas the opposite pattern was observed in DAIC-WOZ. For example, AU26_pk_int showed an effect size of −0.658 in the Korean cohort (happy condition, PHQ-8 definition) but +0.267 in DAIC-WOZ. Thus, the peak-interval features that produced the strongest within-cohort discrimination (Section IV-C) reversed direction in the external dataset. Recording-length adjustment was applied to the core AU effects (Section IV-E) but not to peak features, so the contribution of recording length to this reversal was not separately estimated.

The preservation of non-peak features was supported statistically. Directional agreement significantly exceeded chance for non-peak features (binomial test: happy 18/20, p = 2.0 × 10^-4^; unhappy 32/39, p = 3.5 × 10^-5^). Their effect sizes were also positively correlated between datasets (happy condition: Pearson r = 0.58, p = 0.007), whereas peak-related features showed a significant negative correlation (Spearman ρ = −0.41, p = 0.03), confirming the systematic reversal. At the AU level, AU06 exhibited the greatest robustness, with directional agreement in 12/13 features in the happy condition and 11/11 in the unhappy condition. Because features derived from the same AU are not statistically independent, the binomial test should be regarded as supportive rather than definitive; the principal evidence for transferability comes from the consistent AU-level agreement and the cross-dataset effect-size correlations.

Together, these findings demonstrate that the features producing the largest within-cohort effects were not necessarily those that generalized across datasets. In contrast, AU06-centered temporal features showed modest effect sizes but consistently preserved their direction across cohorts with substantial differences in population characteristics and recording protocols, supporting their potential as transferable facial biomarkers of depression.

### E. Robustness Analysis

We evaluated the robustness of the identified biomarkers with respect to recording length and demographic characteristics.

As shown in Fig. 3, adjustment for recording length preserved the direction of the core AU effects. Within recording-length tertiles, the directions of the early-, middle-, and late-session effects remained consistent for 18/18 features in the happy condition and 17/18 features in the unhappy condition. AU06 and AU07 exhibited even larger effect sizes in the longer speech segments.

**Fig. 3.**
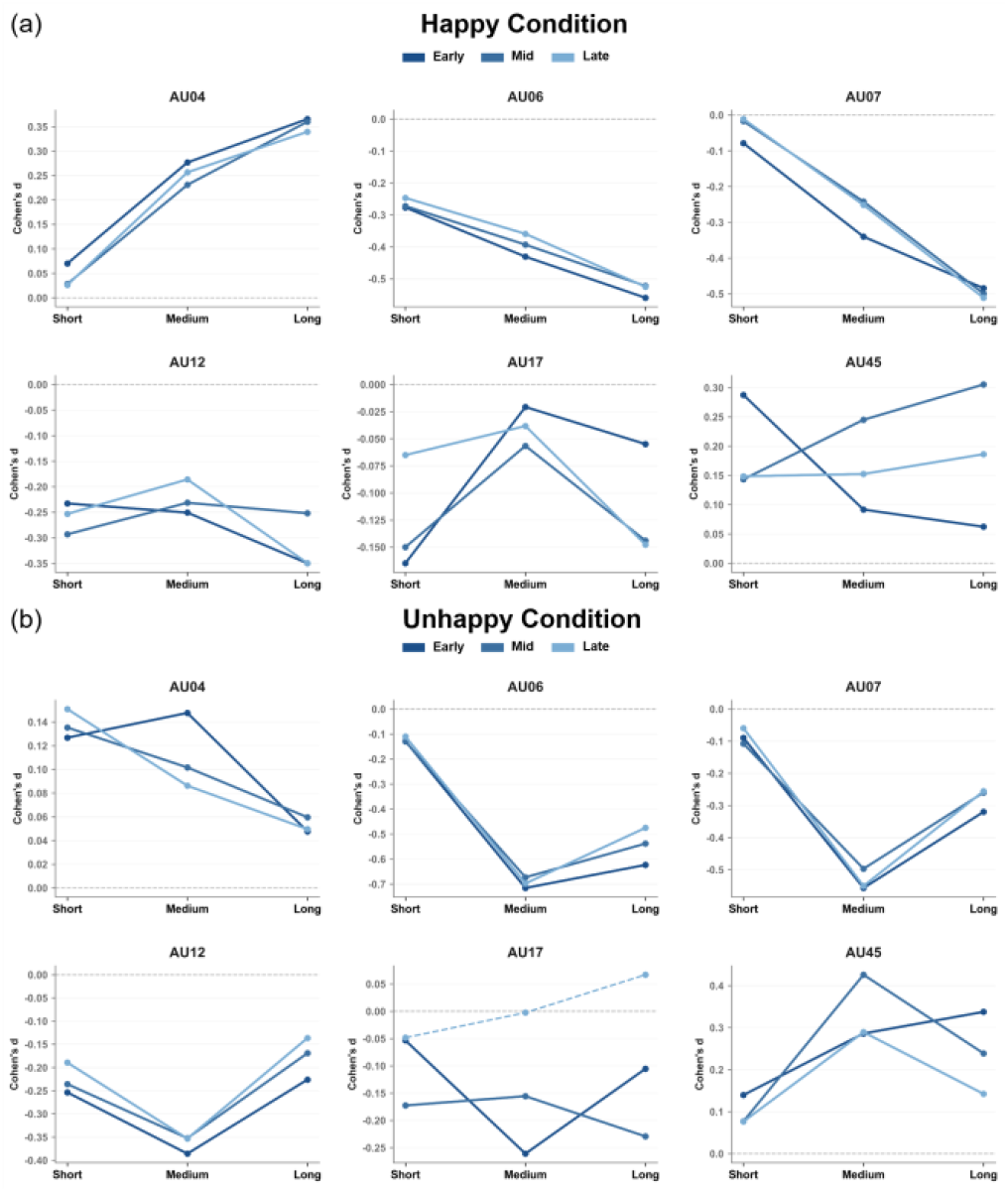
Stability of core AU effects across recording-length tertiles. Cohen’s d values for representative core AUs across short, medium, and long recording-length tertiles in the happy and unhappy (b) conditions. Most core AU effects preserve their direction across tertiles, indicating that the observed depression-related differences are not driven by recording length.

The identified biomarkers also remained stable across demographic subgroups. Directional consistency exceeded approximately 80% across sex, age (65-year cutoff), and speech-length subgroups (Supplementary Fig. S2). Inconsistent directions were largely confined to peak features and the older subgroup (Supplementary Table S4). Peak features accounted for six of the seven largest between-subgroup differences in effect size, and three peak-interval features reversed direction in the long-recording tertile, so their instability appears within the source cohort as well as across datasets. Because only 44 depressed participants were aged 65 or older, effects in that subgroup were interpreted cautiously. Overall, 131 features remained significant in both emotional conditions with concordant directions, led by AU12 (21 features) and AU06 (17).

Using these robust features, a model combining AU06 temporal and smile measures reached a 5-fold cross-validated AUROC of 0.61 in the happy condition and 0.62 in the unhappy condition, whereas a peak-only model was far less consistent across conditions (0.70 versus 0.57). After adjustment for age, sex, and speech length, au06_given_au12 and smile quality remained associated with depression in both conditions (p < 0.05), while the intensity-level measures were attenuated: AU06_mean was not significant in either condition, and AU06_early only in the happy condition (p = 0.031 happy, 0.061 unhappy). Detailed results are in Supplementary Table S5.

### F. Smile Representation Analysis

Fig. 4 shows that the depressed group exhibited a lower proportion of Duchenne, or genuine, smiles (both approximately d = −0.29) and a higher proportion of non-Duchenne, mouth-only smiles (happy d = +0.31, unhappy d = +0.21). This resulted in lower smile quality in the depressed group (happy d = −0.31, unhappy d = −0.26). The depressed group did not smile less overall; rather, the qualitative composition of smiling differed (Supplementary Fig. S3).

**Fig. 4.**
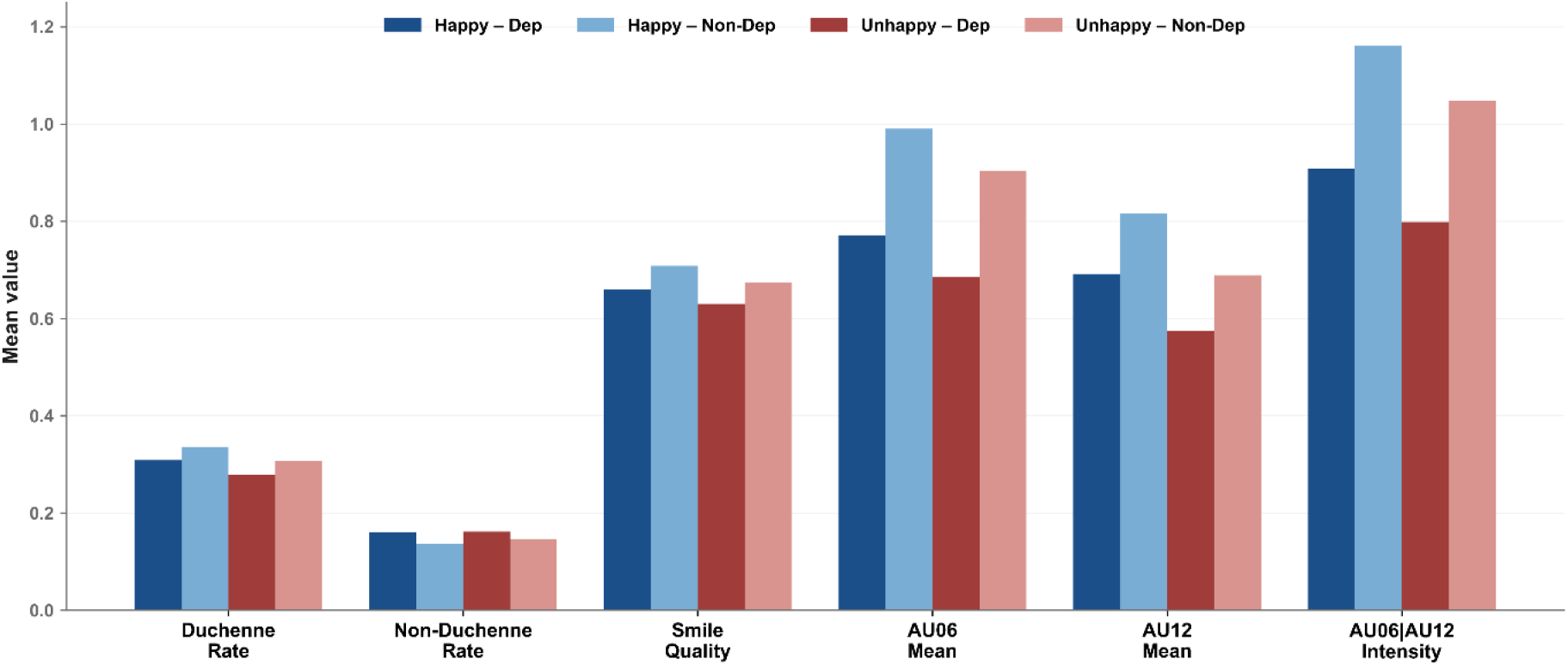
Smile composition in depression. Mean values of Duchenne rate, non-Duchenne rate, smile quality, AU06 mean intensity, AU12 mean intensity, and AU06 intensity conditioned on AU12 activation, stratified by depression status and emotional condition.

The strongest smile-related measure was au06_given_au12, defined as the intensity of AU06 in frames where AU12 was active. This metric captures the extent to which eye activity accompanies mouth movement after controlling for smile frequency. It was lower in the depressed group (happy d = −0.410, unhappy d = −0.416) and showed the largest effect size among the 17 smile-related measures. The direction of this effect remained consistent across sex, age, and speech-length subgroups, although it was attenuated in the older subgroup (Supplementary Fig. S4).

Temporal trajectories of genuine smiling are presented in Fig. 5, which shows group mean Duchenne rates across session thirds; the corresponding effect sizes are provided in Supplementary Fig. S5. Group differences were largest during the early segment of the session and progressively diminished over time. In the happy condition, the effect size of the Duchenne rate decreased from d = −0.222 in the early segment to d = −0.072 in the late segment, with a similar pattern observed in the unhappy condition. Because the temporal segments represent relative thirds of valid frames, they indicate relative position within a session rather than absolute elapsed time. Associations between individual PHQ-9 symptom items and AU features are summarized in Supplementary Table S6.

**Fig. 5.**
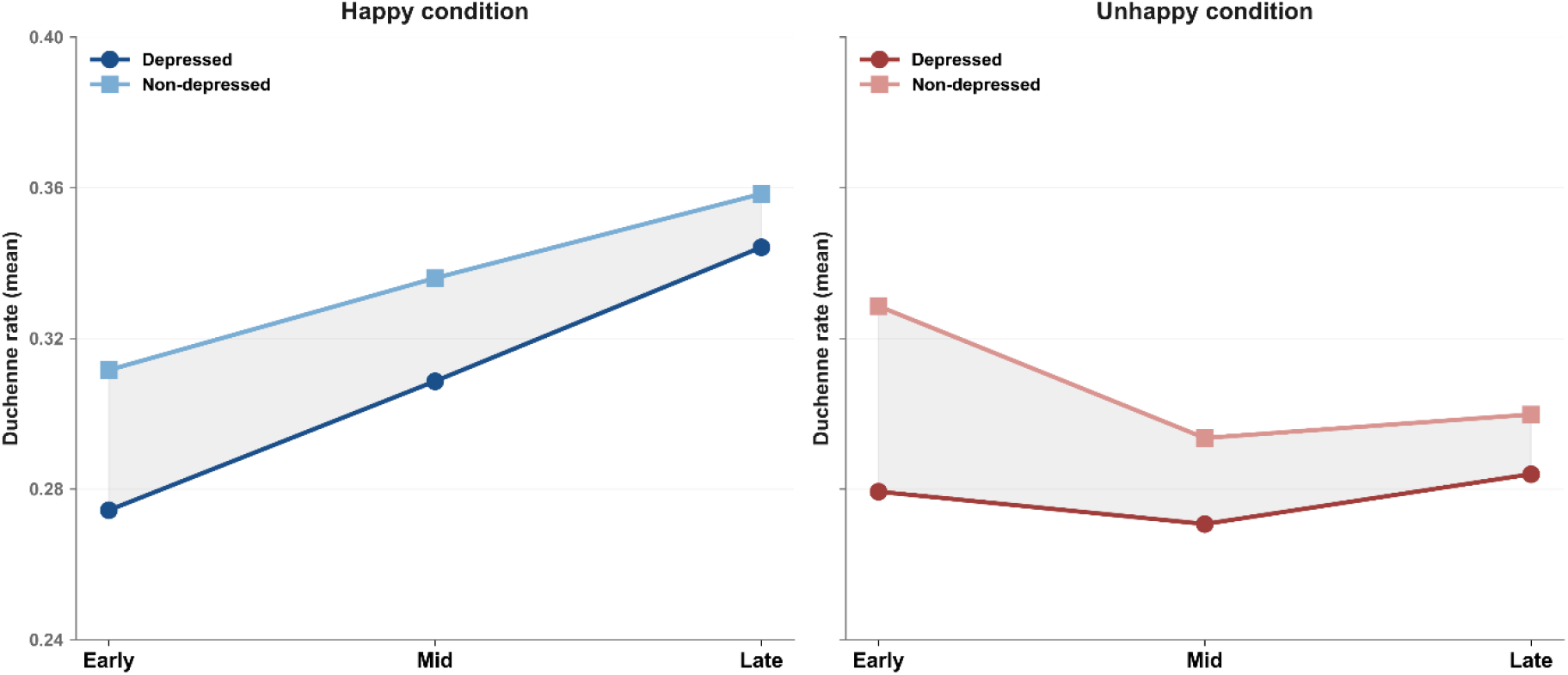
Temporal course of genuine smiling. Group means of Duchenne rate across relative session thirds (early, mid, late) in the happy and unhappy conditions. Group differences are largest in the early segment and become smaller in later segments. Segments represent relative thirds of valid frames rather than absolute elapsed time.

### G. Integrated Biomarker Summary

We integrated multiple validation criteria to identify the most reliable depression-related facial biomarkers. These criteria included Bonferroni significance, FDR significance with concordant direction across emotional conditions, directional agreement with DAIC-WOZ, robustness to recording-length adjustment, and stability across recording-length tertiles (Fig. 6).

**Fig. 6.**
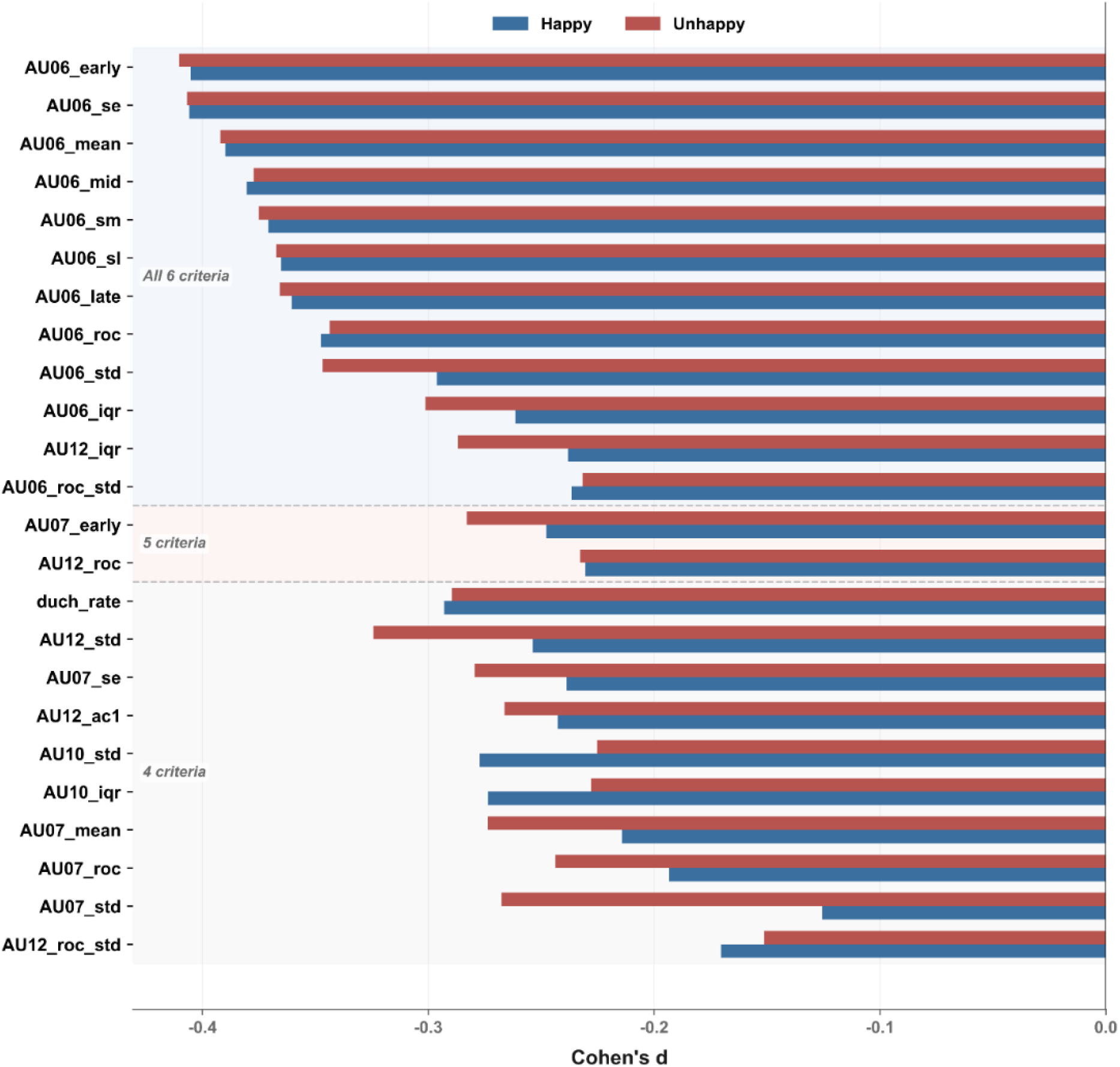
AU06 as the most consistent biomarker across validation criteria. Features meeting multiple validation criteria, including Bonferroni significance, FDR significance with concordant direction across emotional conditions, directional agreement with DAIC-WOZ, robustness to recording-length adjustment, and stability across recording-length tertiles. Features are ordered by the number of criteria satisfied. AU06-derived features dominate the fully validated set.

When all criteria were applied, 12 features satisfied every requirement, and 11 of them were derived from AU06, including AU06_early, AU06_mean, and AU06_roc. Whereas the robust core defined by within-cohort criteria alone was led by AU12 (Section IV-E), adding cross-dataset directional agreement progressively narrowed the validated set to AU06. This indicates that AU06 carried the facial signal that was both reproducible within the Korean cohort and transferable to the external dataset.

The convergence between the temporal-feature analysis and the smile representation analysis was also centered on AU06. The features that generalized most consistently across datasets were predominantly AU06-derived temporal and variability measures, whereas peak-related features, despite large within-cohort effects, did not transfer (Section IV-D). In parallel, the strongest smile-related measure, au06_given_au12, depended on AU06 through its coactivation with AU12 (Section IV-F). Thus, both analytic axes identified AU06 as the most consistently validated facial action unit associated with depressive symptoms, suggesting that AU06 carries depression-related information both as a standalone temporal feature and as the eye-region component of Duchenne-type smiling.

## V. DISCUSSION

### A. Principal Findings

In a Korean cohort of 2,608 participants, we analyzed 568 temporal features of 17 AUs together with the qualitative composition of smiling, and externally validated the markers in the US DAIC-WOZ dataset [18], which differs along five axes including race and interview task. This study makes three contributions. First, it reproduces, in a large cohort, AU-depression findings that have until now rested on small samples [6, 7]. Second, it tests whether the core markers preserve the direction of their effect in a heterogeneous external dataset, thereby distinguishing the types of features that generalize from those that do not. Third, through temporal analysis and an analysis of smile authenticity, it identifies markers that aggregate statistics fail to capture and examines their mechanistic meaning.

### B. Reproduction in a Large Cohort

Most prior AU-depression studies involved tens to a few hundred participants [6, 7, 12]. In 2,608 participants, we confirmed reduced facial activity in the depressed group, centered on AU06 (cheek raiser). The mean intensity of AU06 was lower in the depressed group in both emotional conditions, and AU12 (lip corner puller) was in the same direction. This is consistent with prior reports that positive-emotion-related facial expression is reduced in depression [7, 9]. That the direction of these effects reproduced consistently in a large sample indicates that the findings are not an artifact of small samples. Recent AU-based depression models have likewise identified AU06 and AU12 as features of the non-depressed state, but on much smaller samples. An interpretable LSTM model with integrated-gradient explanations associated AU06, AU12, AU14, and AU25 with non-depression and AU26, AU20, and AU07 with depression in 474 interview videos (134 depressed) [11], and a two-stream model linking facial and body expression reported group differences concentrated in AU06, AU07, AU10, AU12, and AU14 in 156 participants [31]. Our cohort is larger by an order of magnitude, and the consistent direction of the AU06 effect at this scale strengthens the evidence that the finding is not sample-specific. Age warrants comment, as AU06 acts on the periocular region and its automatic estimation is sensitive to wrinkling. The depressed group was younger by a median of 13 years, so age-related bias would tend to raise rather than lower measured AU06 in that group; the observed effect runs opposite to this bias.

### C. External Generalization and Generalizable Feature Types

Most AU-depression studies are based on a single dataset, typically DAIC-WOZ or its extension E-DAIC [18, 23, 32], and report within-dataset classification performance. Within-dataset performance, however, does not guarantee the validity of a marker. Many DAIC-WOZ depression classifiers have been shown to learn cues of general psychological distress rather than depression-specific signals, casting doubt on their validity [21], and facial-expression datasets are known to carry collection-related bias that hinders cross-dataset generalization [33]. More broadly, poor cross-domain transfer is a known weakness of automated facial analysis. A review of 39 studies found that cross-domain performance consistently falls short of within-domain performance, and that this gap has been obscured by the common practice of reporting aggregate rather than label-specific results [34]. When AU detectors are trained on one database and tested on another, detection accuracy usually drops, often below the level needed for behavioral research, and the size of this drop varies from one action unit to the next; the same work showed, through occlusion analysis, that the facial regions a detector relies on within-domain are more local than those it uses cross-domain [35]. These studies measured the detection accuracy of AU classifiers. Our question is different: holding AU extraction fixed, we ask whether the direction of an AU-derived depression marker is preserved across datasets, and which feature types preserve it. The contrast between local within-domain cues and more diffuse cross-domain cues parallels our own finding, in which the peak-interval features that give the strongest within-dataset signal are exactly the ones that fail to transfer.

The same limitation on cross-dataset transfer recurs in adjacent modalities. In facial expression recognition, cross-dataset accuracy drops sharply with the choice of source and target, and local, fine-grained features transfer better than holistic ones [36]; in speech, cross-corpus emotion models degrade under shifts in recording conditions, elicitation, and speaker demographics [37]. These studies recover performance by learning domain-invariant representations through adversarial training. We take the opposite approach: without any adaptation, we ask which existing AU-derived markers are already transferable, and identify them by the directional consistency of their effects. Because temporal dynamics preserved their direction even though the two datasets differed greatly in absolute scale, this consistency reflects information carried by the shape of the signal rather than by its intensity.

Our external dataset, DAIC-WOZ, differs from the Korean cohort along five axes at once: race, language, interview task, recording length, and recording setting. Because these axes differ simultaneously, the comparison is not a simple replication in an additional dataset but a stress test of generalizability. From a domain-adaptation standpoint, direction preserved under such distant conditions implies transfer to more similar datasets; the strength of this validation therefore rests on the heterogeneity of the external data rather than the number of datasets used. Against this benchmark, the feature types split cleanly. Temporal, central-tendency, and dynamics features preserved their direction, whereas the peak-interval features that gave the strongest within-cohort signal reversed. A plausible source of the reversal is the interview task itself: the Korean recordings are a few minutes of structured speech, whereas DAIC-WOZ is a longer free interview [18], which would shape the distribution of peaks differently. The same account extends to the activation-threshold features, the other category that transferred poorly: both peak and activation features are defined by event timing relative to a within-session threshold, whereas the features that transferred describe the level and shape of the signal. That the strongest within-cohort signal is the one that fails externally shows directly why within-dataset effect size cannot guide feature selection.

### D. Temporal Markers and Qualitative Change in Smiling

Unlike prior work that relied on aggregate statistics [6, 12], this study identified markers in temporal features. Most features that passed all validation criteria were AU06 temporal features, which carry information not contained in mean values, such as the rate of change and early-session activity. The within-cohort robust set, by contrast, was led by AU12 (Section IV-E). AU12 is under voluntary control and its display responds to the recording setting, whereas AU06 is not readily produced on demand; the narrowing from AU12 to AU06 under external validation follows this asymmetry.

Several recent models have similarly argued that the temporal course of facial expression, rather than static intensity, carries the depression signal: a bidirectional RNN model reported that the expression curves of depressed individuals are smoother over time than those of non-depressed individuals [38], and a multimodal model integrating large language models with facial features used a bidirectional temporal architecture over per-frame AUs [32]. These models, however, were each developed and tested within a single dataset; our results add that only a subset of such temporal features preserves its direction across datasets. In the analysis of smile quality, the depressed group showed fewer Duchenne smiles, more non-Duchenne smiles, and lower smile quality in both conditions. The classical claim that the Duchenne smile is associated with genuine positive emotion [15, 16] was thus reproduced at scale in a depression context. The largest effect appeared in au06_given_au12, the degree to which eye activity accompanies the mouth during smiling. The qualitative composition of smiling was more strongly linked to depression than the average frequency of smiling. A parallel pattern has been reported in speech, where, among several emotional dimensions, only valence transferred across corpora and languages as a depression indicator [26], echoing our finding that the signal concentrates in a single component, the eye activity captured by AU06, rather than in the overall amount of expression.

It has been argued that AU06 may be an artifact of smile intensity rather than an indicator of genuine positive emotion [39]. Our au06_given_au12 compares AU06 only in frames where AU12 is active, thereby controlling for the occurrence of smiling itself. That this measure remained significantly associated with depression after adjustment for age, sex, and speech length, whereas the intensity-level measures did not, indicates that the decoupling of the eyes and mouth is a signal distinct from a general blunting of expression. Given that AU06 is difficult to produce voluntarily whereas AU12 can be generated deliberately [15], this decoupling leads to the hypothesis of a dissociation between spontaneous emotional processing and voluntary motor expression. This provides a mechanism-oriented hypothesis that can be tested in future work combining neuroimaging or physiological measurement.

This dissociation maps onto a well-established neuroanatomical distinction between two pathways that both converge on the facial nucleus but originate separately: voluntary expressions are driven by the motor cortex via the pyramidal tract, whereas genuine emotional expressions arise from subcortical and limbic structures, including the amygdala, and reach the facial nucleus through extrapyramidal pathways [40–42]. Because the pyramidal system exerts only imperfect control over the dynamics of genuine expression, the eye– mouth decoupling captured by au06_given_au12 is consistent with reduced engagement of the extrapyramidal, emotion-driven pathway relative to preserved voluntary mouth movement, rather than a global motor deficit. We frame this as a falsifiable behavioral prediction rather than a demonstrated neural mechanism: if the decoupling indexes extrapyramidal involvement, it should dissociate from voluntary AU12 control under direct neural or physiological measurement, a prediction the present behavioral data cannot test but can motivate. This interpretation is consistent with the clinical dissociation between emotional and volitional facial paresis, in which focal lesions selectively impair one pathway while sparing the other [40]. The decoupling may thus represent a candidate behavioral correlate of emotion-specific pathway involvement.

That the increase in non-Duchenne smiling was larger in the happy condition is consistent with meta-analytic findings that emotional reactivity in depression is reduced to both positive and negative stimuli, with a greater reduction for positive stimuli (emotion context insensitivity) [43]. Our results show that this reduction appears not merely as smiling less but as a changed composition of smiling. Temporally, the group difference was largest early in the session and narrowed toward the end. Given that social context influences the display of Duchenne smiles [44], the early group difference may partly reflect a difference in social expression. Because the temporal segments are relative positions rather than absolute time, and depressed participants spoke for shorter periods, this interpretation requires caution.

### E. Symptom-level Associations

Correlations between individual PHQ-9 items and AU features were generally weak and not concentrated on any particular symptom. The facial markers appear to reflect a broad behavioral correlate of depression rather than a direct indicator of a single symptom. Fatigue, sleep, and depressed mood ranked highest in both conditions, and in the unhappy condition AU06 temporal features showed the strongest correlations regardless of symptom type, so that the core marker converged even at the symptom level. Given that the PHQ-9 total compresses heterogeneous symptoms into one score [30], the observation of a consistent signal even at the item level is meaningful.

### F. Implications: a Transferable Phenotype and Clinical Use

Our results suggest that AU features are not only a learnable signal but may also serve as a shareable input that can be distributed across institutions and countries without transferring video. Facial video is itself biometric identifying information, and its re-identification risk and storage demands form a practical bottleneck for multi-site training and external validation. AU intensity time-series, by contrast, are low-dimensional signals extracted from video; they carry lower re-identification risk than the source video and are lightweight, making them comparatively easy to share. This is not merely hypothetical: a widely used resource such as the Extended DAIC-WOZ challenge provides standardized OpenFace-derived action-unit features alongside pose and gaze descriptors as a common visual representation [45], and recent work continues to build depression models directly on these released features [46].

Identifying which AU features are consistently associated with depression across datasets is therefore directly tied to identifying transferable inputs that do not require the video itself. Here, temporal-dynamic features preserved their direction in an external dataset differing along all five axes, suggesting that such features are candidates for privacy-preserving sharing and learning across institutions. This implication, however, rests on three conditions: the extraction procedure must be standardized across sites, since tool version and settings affect feature values and transferability [10]; the present validation extends only to direction-level generalization, not to the transfer of a trained model’s predictive performance; and the re-identification risk of AU features, though lower than that of video, is not zero and requires assessment before sharing. The study should therefore be read not as demonstrating the feasibility of AU-based sharing but as empirically identifying the type of transferable features on which such an approach would have to rely.

These findings also point to a practical role for facial analysis as an auxiliary signal in depression screening. Because the classification performance falls short of standalone diagnosis, it is best used alongside other modalities such as self-report or speech and language [47]. The operational message is concrete: in video-based screening, weighting the temporal dynamics centered on AU06 and AU12, rather than the absolute magnitude or peak structure of expression, is more robust across datasets. In the same vein, when observing depression in a clinical interview, it may be useful to attend to whether the eyes follow the mouth during smiling.

### G. Limitations

The core effects were robust to adjustment for speech length, consistent within length tertiles, and largely preserved across sex and age subgroups, but several limitations remain. First, the cross-sectional design precludes causal inference. Second, depression status relied on the PHQ-9 self-report, which carries misclassification risk relative to a structured clinical interview [30]. As reported in a recent meta-analysis, however, the PHQ-9 has become a widely used approach for scalable depression labeling in large-scale and machine-learning studies; that meta-analysis treated the use of validated, standardized measures such as the PHQ as a marker of study quality [48]. Third, the external validation used a single dataset (DAIC-WOZ), and because the interview structure differed, the design cannot separate racial from task sources of the peak-feature reversal; validation in two or more independent datasets under an identical protocol is needed. Fourth, peak features were not adjusted for recording length, so protocol and length effects cannot be separated. Fifth, the depressed subgroup aged 65 or older was small, so those results warrant caution. Sixth, OpenFace [10] AU recognition is imperfect and its performance on East Asian faces may differ.

## VI. CONCLUSION

Research on facial markers of depression has largely been evaluated by within-dataset classification performance. This study shows that such performance does not guarantee transferability: the strongest within-dataset signal, the peak-interval features, reversed in direction in a dataset differing in race, language, task, and recording setting, whereas AU06 and slower temporal features, despite their modest effect sizes, preserved their direction. Among these, what survived adjustment for age, sex, and speech length was not the level of AU06 but its coupling to AU12 during smiling. We formalize the implication as a directional-consistency screening criterion: a feature qualifies as a transfer candidate only if the sign of its effect is preserved in a highly heterogeneous external dataset, independent of its within-dataset effect size. Applied here, the criterion retains temporal-segment, central-tendency, and dynamics features while rejecting peak-structure features, offering a reusable selection rule for AU-derived inputs rather than a finding specific to this cohort. Because AU time-series are lower-dimensional representations with lower direct identifiability than raw video, features selected this way may support privacy-conscious multi-site research, provided that re-identification risk is formally assessed. Clinically, these markers are best used not as a standalone screening tool but as an auxiliary signal. Future work should validate the transfer of these features across multiple independent datasets and, beyond direction-level agreement, determine whether multi-site learning that shares only AUs achieves useful predictive performance.

## Data Availability

The data and code used in this study are not publicly available. The facial video recordings and derived data were collected under institutional review board protocols that do not permit public release because of privacy and ethical considerations associated with identifiable facial data from human participants. De-identified derived data and analysis code may be made available from the corresponding authors upon reasonable request, subject to institutional review board approval and a data use agreement. The external validation dataset, DAIC-WOZ, is available from its original distributors under their respective access procedures.

## Supplementary Tables

**Supplementary Table S1.** Facial Action Units Included in the Analysis.

| **AU** | **FACS Name** | **Facial Movement** | **Common Emotional Interpretation** |
| --- | --- | --- | --- |
| **AU01** | Inner Brow Raiser | Raises the inner portion of the eyebrows | Sadness, concern |
| **AU02** | Outer Brow Raiser | Raises the outer portion of the eyebrows | Surprise, fear |
| **AU04** | Brow Lowerer | Lowers and draws the eyebrows together | Negative affect; hypothesized to increase in depression |
| **AU05** | Upper Lid Raiser | Raises the upper eyelids, widening the eyes | Surprise, arousal |
| **AU06** | Cheek Raiser | Raises the cheeks and creates wrinkles around the eyes | Eye component of a Duchenne smile |
| **AU07** | Lid Tightener | Tightens the eyelids | Tension, negative affect |
| **AU09** | Nose Wrinkler | Wrinkles the nose | Disgust |
| **AU10** | Upper Lip Raiser | Raises the upper lip | Disgust, contempt |
| **AU12** | Lip Corner Puller | Pulls the lip corners upward (smile) | Positive affect; mouth component of smiling |
| **AU14** | Dimpler | Tightens the lip corners laterally | Contempt, restrained emotion |
| **AU15** | Lip Corner Depressor | Pulls the lip corners downward | Characteristic marker of sadness |
| **AU17** | Chin Raiser | Raises the chin and pushes up the lower lip | Doubt, displeasure |
| **AU20** | Lip Stretcher | Stretches the lips horizontally | Fear |
| **AU23** | Lip Tightener | Presses and tightens the lips together | Anger, tension |
| **AU25** | Lips Part | Parts the lips | Often accompanies speech |
| **AU26** | Jaw Drop | Lowers the jaw and opens the mouth | Surprise, speech-related movement |
| **AU45** | Blink | Eye blink | Arousal, cognitive load |

*The table lists the 17 facial action units (AUs) extracted using OpenFace and analyzed in this study. Each AU is described according to the Facial Action Coding System (FACS), together with its corresponding facial movement and commonly reported emotional interpretation. Emotional interpretations are provided for descriptive purposes and should not be regarded as one-to-one mappings between individual AUs and specific emotions.*

**Supplementary Table S2.** Temporal and Coordination Features Extracted from AU Time Series.

| Feature name | Category | Definition |
| --- | --- | --- |
| **AU_mean** | Central tendency | Mean AU intensity across all valid frames |
| **AU_mid** | Central tendency | Mean AU intensity in the middle third of the session |
| **AU_early** | Temporal segment | Mean AU intensity in the first third of the session |
| **AU_late** | Temporal segment | Mean AU intensity in the final third of the session |
| **AU_se** | Temporal segment | Mean AU intensity in the first two chunks after dividing the session into five equal chunks |
| **AU_sm** | Temporal segment | Mean AU intensity in the middle chunk after dividing the session into five equal chunks |
| **AU_sl** | Temporal segment | Mean AU intensity in the final two chunks after dividing the session into five equal chunks |
| **AU_slope** | Shape/trend | Linear slope of AU intensity across frames |
| **AU_le** | Shape/trend | Difference in mean AU intensity between the late and early thirds |
| **AU_sle** | Shape/trend | Difference in mean AU intensity between the final and first segments in the five-chunk division |
| **AU_ss** | Shape/trend | Linear slope of AU intensity across the five session chunks |
| **AU_std** | Variability | Standard deviation of AU intensity |
| **AU_iqr** | Variability | Interquartile range of AU intensity |
| **AU_skew** | Variability | Skewness of the AU intensity distribution |
| **AU_kurt** | Variability | Kurtosis of the AU intensity distribution |
| **AU_roc** | Dynamics | Mean absolute frame-to-frame change in AU intensity |
| **AU_roc_std** | Dynamics | Standard deviation of frame-to-frame changes in AU intensity |
| **AU_ac1** | Dynamics | Autocorrelation of AU intensity at a lag of 1 frame |
| **AU_ac5** | Dynamics | Autocorrelation of AU intensity at a lag of 5 frames |
| **AU_ac10** | Dynamics | Autocorrelation of AU intensity at a lag of 10 frames |
| **AU_burst_n** | Burst pattern | Number of bursts exceeding the 75th percentile of within-session AU intensity |
| **AU_burst_dur** | Burst pattern | Mean duration of AU bursts |
| **AU_burst_max** | Burst pattern | Maximum duration of AU bursts |
| **AU_ibi_mean** | Burst pattern | Mean interval between the onset of successive AU bursts |
| **AU_ibi_cv** | Burst pattern | Coefficient of variation of inter-burst intervals |
| **AU_act_n** | Activation pattern | Number of activation episodes above the within-session median |
| **AU_act_rate** | Activation pattern | Proportion of frames in which the AU was active |
| **AU_on2off** | Activation pattern | Transition rate from active to inactive states |
| **AU_off2on** | Activation pattern | Transition rate from inactive to active states |
| **AU_npk** | Peak structure | Number of detected peaks in the AU intensity signal |
| **AU_pk_int** | Peak structure | Mean interval between successive AU intensity peaks |
| **AU_pk_cv** | Peak structure | Coefficient of variation of inter-peak intervals |
| **pair_corr** | AU-pair coordination | Within-frame correlation between two AU intensity time series |
| **pair_bestlag** | AU-pair coordination | Time lag at which the correlation between two AU time series was maximal |
| **coact_mean** | Duchenne/coactivation | Mean number of simultaneously active AUs per frame |
| **duch_rate** | Duchenne/coactivation | Proportion of frames with simultaneous activation of AU06 and AU12 |
| **duch_dur** | Duchenne/coactivation | Mean duration of Duchenne smile episodes |
| **duch_n** | Duchenne/coactivation | Number of Duchenne smile episodes |

*For each of the 17 AUs, 32 temporal features were extracted from the frame-wise intensity signal, including measures of central tendency, temporal segments, variability, dynamics, burst patterns, activation patterns, and peak structure. In addition, two coordination features were computed for each predefined AU pair, and four global facial-coordination features were derived. Together, these features yielded a total of 568 variables used for statistical analysis.*

**Supplementary Table S3.**
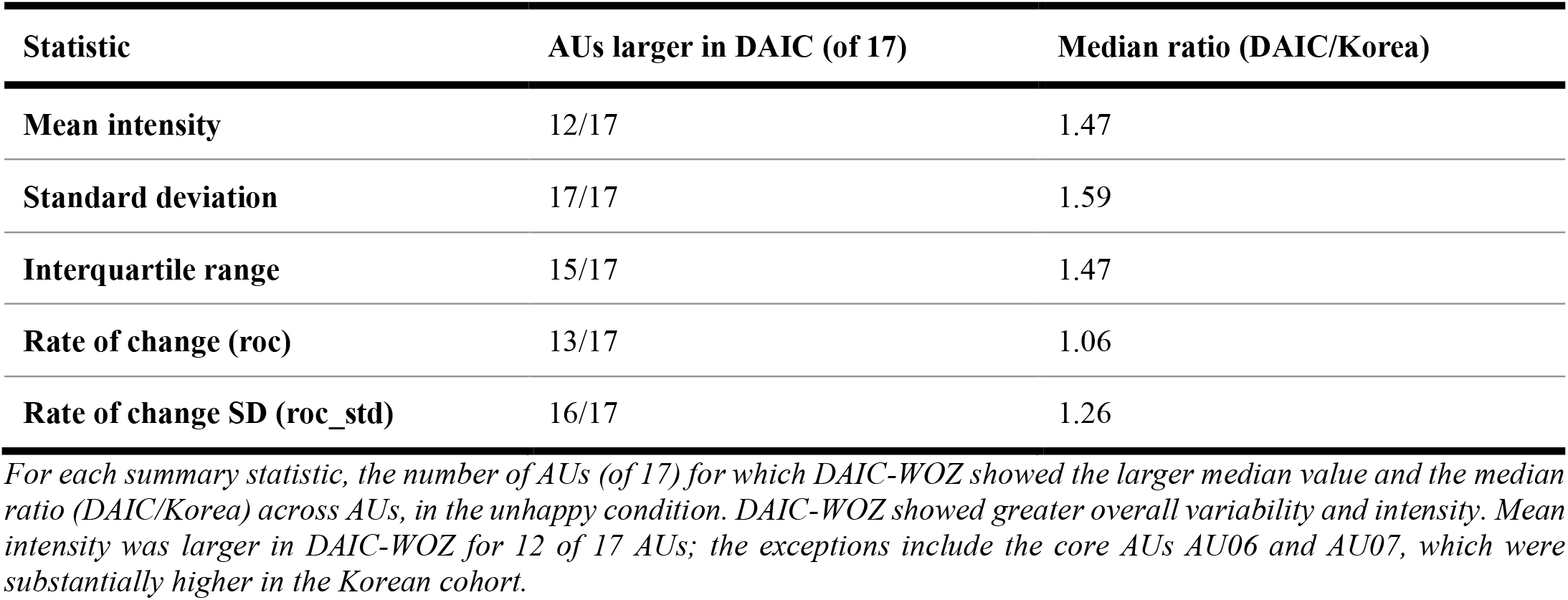
Comparison of AU summary statistics between the Korean cohort and DAIC-WOZ.

| Statistic | AUs larger in DAIC (of 17) | Median ratio (DAIC/Korea) |
| --- | --- | --- |
| Mean intensity | 12/17 | 1.47 |
| Standard deviation | 17/17 | 1.59 |
| Interquartile range | 15/17 | 1.47 |
| Rate of change (roc) | 13/17 | 1.06 |
| Rate of change SD (roc_std) | 16/17 | 1.26 |

*For each summary statistic, the number of AUs (of 17) for which DAIC-WOZ showed the larger median value and the median ratio (DAIC/Korea) across AUs, in the unhappy condition. DAIC-WOZ showed greater overall variability and intensity. Mean intensity was larger in DAIC-WOZ for 12 of 17 AUs; the exceptions include the core AUs AU06 and AU07, which were substantially higher in the Korean cohort.*

**Supplementary Table S4.** Sign Reversals and Magnitude Differences Across Demographic Subgroups.

reversals (subgroup d opposite to overall, $|d| > 0.1$ ):
| Feature | Condition | Subgroup | Overall d | Subgroup d |
| --- | --- | --- | --- | --- |
| AU01_pk_int | unhappy | Len=Long | -0.159 | +0.261 |
| AU02_pk_int | unhappy | Len=Long | -0.163 | +0.267 |
| AU05_pk_int | unhappy | Len=Long | -0.125 | +0.279 |
| AU07_early | unhappy | Age≥65 | -0.285 | +0.214 |
| AU07_mean | unhappy | Age≥65 | -0.270 | +0.165 |
| AU07_se | unhappy | Age≥65 | -0.281 | +0.213 |
| AU07_roc | unhappy | Age≥65 | -0.251 | +0.137 |
| AU09_burst_n | unhappy | Age≥65 | +0.318 | -0.215 |
| AU09_npk | happy | Len=Long | -0.322 | +0.112 |

Largest magnitude differences between subgroups ( $|\Delta d|$ ):
| Feature | Condition | Contrast | d (group A) | d (group B) | abs $\Delta$ |
| --- | --- | --- | --- | --- | --- |
| AU25_pk_cv | happy | Age (<65 / ≥65) | +0.259 | +1.181 | 0.923 |
| AU06_pk_int | happy | Age (<65 / ≥65) | -0.193 | -0.955 | 0.762 |
| AU20_pk_cv | happy | Age (<65 / ≥65) | +0.163 | +0.728 | 0.564 |
| AU09_ac1 | unhappy | Sex (M / F) | -0.019 | -0.571 | 0.552 |
| AU07_pk_int | happy | Age (<65 / ≥65) | -0.369 | -0.894 | 0.526 |
| AU10_pk_int | happy | Age (<65 / ≥65) | -0.341 | -0.829 | 0.488 |
| AU05_pk_int | happy | Age (<65 / ≥65) | -0.411 | -0.899 | 0.488 |

Features whose effect direction reversed relative to the overall sample, or whose absolute effect size differed substantially between subgroups. Reversals and large magnitude differences were dominated by peak features and the older subgroup, indicating that peak features are context-dependent within the source cohort as well as across datasets. Representative rows are shown; the complete set of 25 sign reversals is available from the corresponding authors upon request. Large magnitude differences in the older subgroup should be interpreted in light of its small depressed sample ( $n=44$ ).

**Supplementary Table S5.**
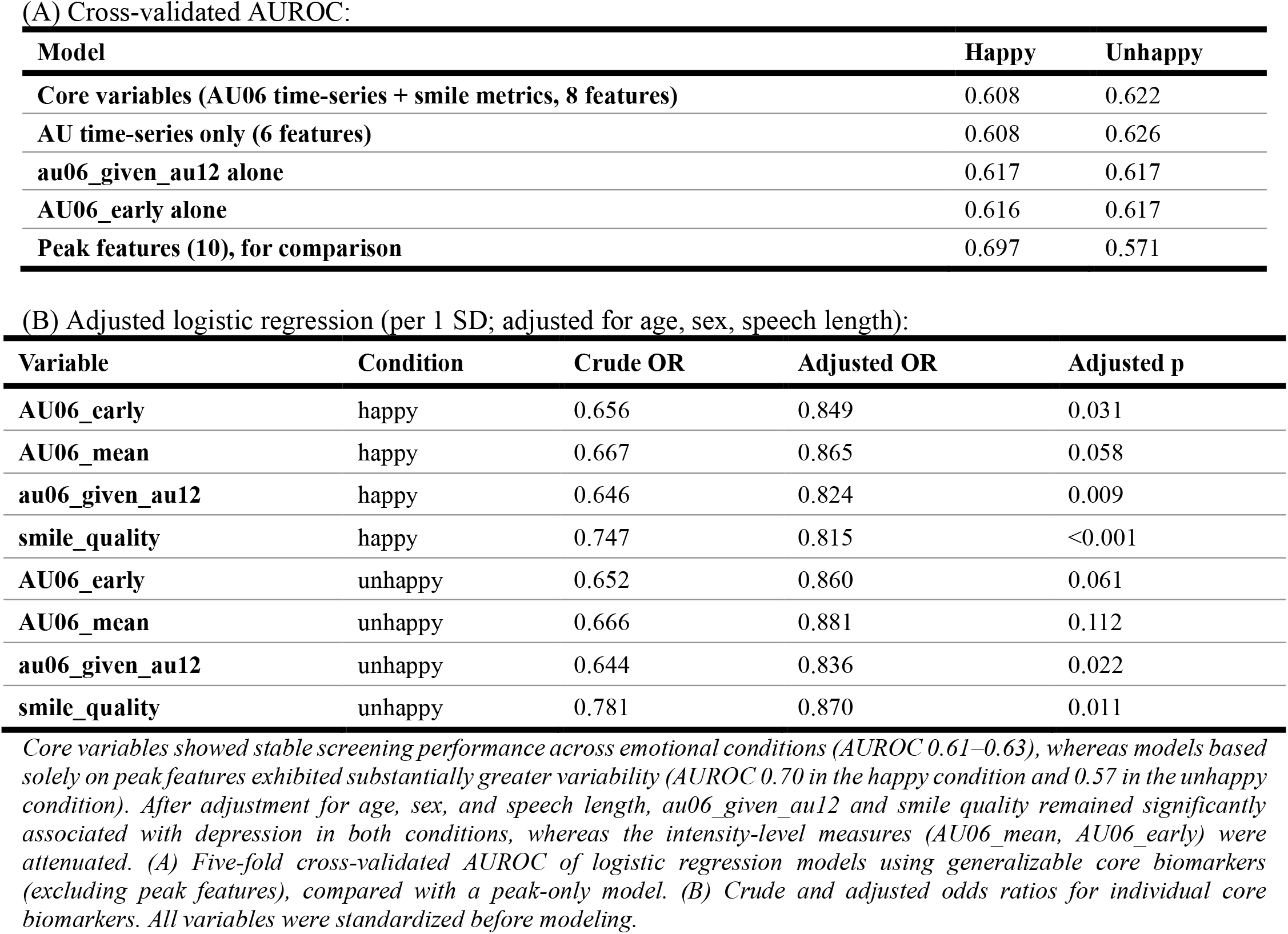
Core-Variable Screening Models and Covariate-Adjusted Associations.

**Supplementary Table S6.** Association Between AU Features and Individual PHQ-9 Symptom Items.

| Rank | PHQ-9 item | Median $\vert \rho\vert $ | Max $\vert \rho\vert $ | n FDR-sig | Top feature ($\rho$) |
| --- | --- | --- | --- | --- | --- |
| 1 | Fatigue | 0.037 | 0.158 | 239 | AU23_npk (-0.158) |
| 2 | Sleep | 0.034 | 0.132 | 203 | AU09_npk (-0.132) |
| 3 | Depressed mood | 0.032 | 0.165 | 190 | AU23_npk (-0.165) |
| 4 | Appetite | 0.028 | 0.132 | 141 | AU23_npk (-0.132) |
| 5 | Anhedonia | 0.026 | 0.145 | 130 | AU23_npk (-0.145) |
| 6 | Worthlessness | 0.024 | 0.146 | 164 | AU23_npk (-0.146) |
| 7 | Concentration | 0.023 | 0.106 | 124 | AU20_pk_cv (0.106) |
| 8 | Suicidal ideation | 0.023 | 0.140 | 107 | AU14_pk_int (-0.140) |
| 9 | Psychomotor | 0.020 | 0.108 | 89 | AU25_roc (-0.108) |

Unhappy condition:
| Rank | PHQ-9 item | Median $\vert \rho\vert $ | Max $\vert \rho\vert $ | n FDR-sig | Top feature ($\rho$) |
| --- | --- | --- | --- | --- | --- |
| 1 | Sleep | 0.033 | 0.125 | 136 | AU06_se (-0.125) |
| 2 | Fatigue | 0.028 | 0.154 | 138 | AU06_early (-0.154) |
| 3 | Depressed mood | 0.027 | 0.154 | 126 | AU06_mean (-0.154) |
| 4 | Appetite | 0.026 | 0.112 | 95 | AU06_se (-0.112) |
| 5 | Worthlessness | 0.024 | 0.121 | 110 | AU06_early (-0.121) |
| 6 | Anhedonia | 0.023 | 0.110 | 69 | AU06_sl (-0.110) |
| 7 | Concentration | 0.021 | 0.077 | 44 | AU45_ac5 (0.077) |
| 8 | Psychomotor | 0.020 | 0.085 | 44 | AU12_std (-0.085) |
| 9 | Suicidal ideation | 0.019 | 0.093 | 16 | AU07_AU45_corr (0.093) |

For each PHQ-9 item, the table reports the median and maximum absolute Spearman correlation ( $|\rho|$ ) across all 568 AU features, the number of FDR-significant features, and the feature showing the strongest association. Items are ranked by median $|\rho|$ within each emotional condition. Correlations were generally modest in magnitude, suggesting that depression-related facial markers are distributed across multiple symptom domains rather than being driven by a single PHQ-9 item.

## Supplementary Figures

**Supplementary Fig. S1.**
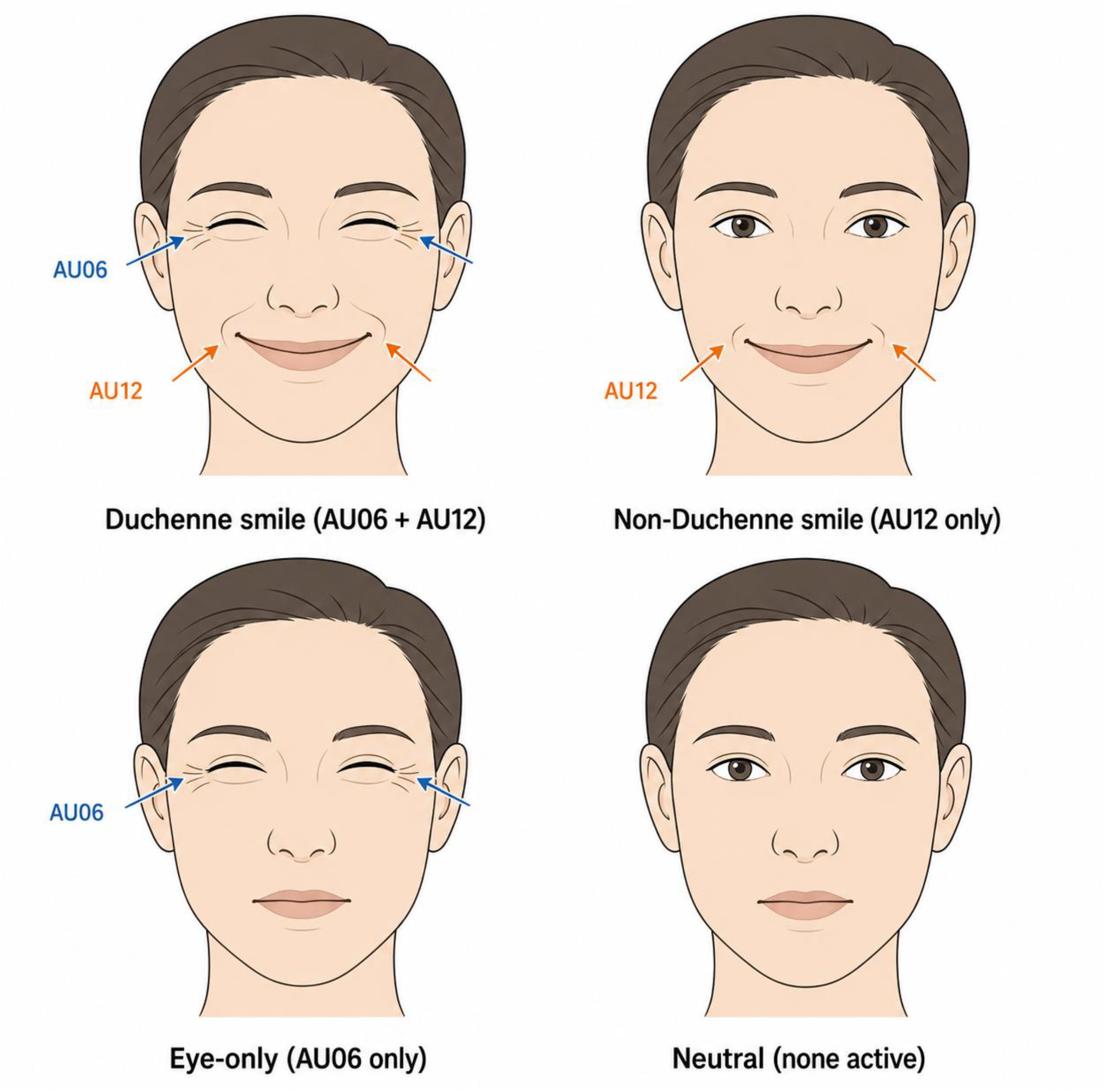
Schematic classification of smile and eye-region expressions based on AU06 and AU12 activation. Four expression types defined by the combination of AU06 (cheek raiser, the involuntary eye component of a genuine smile) and AU12 (lip corner puller). A Duchenne smile shows co-activation of both AU06 and AU12; a non-Duchenne smile shows AU12 activation alone (mouth smiling without eye involvement); an eye-only expression shows AU06 activation alone (eye narrowing without lip corner movement); and a neutral expression shows neither unit active. AU06 accompanies genuine positive affect involuntarily and is difficult to produce on demand, whereas AU12 is under voluntary control, so the co-activation pattern distinguishes genuine from voluntary smiles.

**Supplementary Fig. S2.**
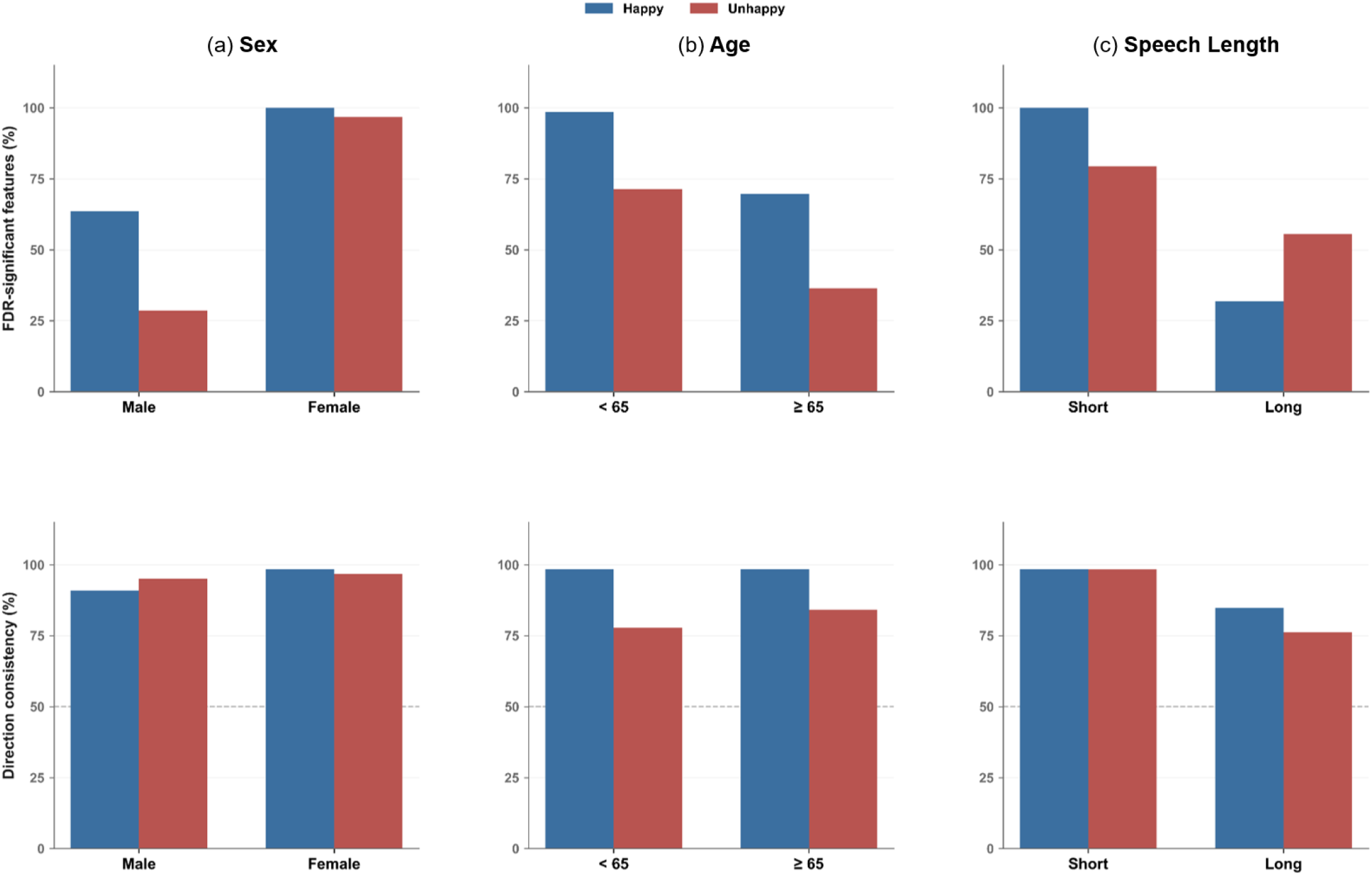
Stability of biomarkers across demographic and speech-length subgroups. (a) Sex, age (65-year cutoff), and (c) speech-length (median split) subgroup analyses. The top row shows the proportion of biomarkers that remained FDR-significant within each subgroup, and the bottom row shows the proportion that preserved the same effect direction (Cohen’s d sign) as observed in the full sample. Results are presented separately for the happy and unhappy conditions. Because the 265 depressed participants were divided across subgroups, smaller strata (particularly depressed participants aged ≥65 years, n = 44) had limited statistical power, and lower significance proportions in these groups should be interpreted with caution.

**Supplementary Fig. S3.**
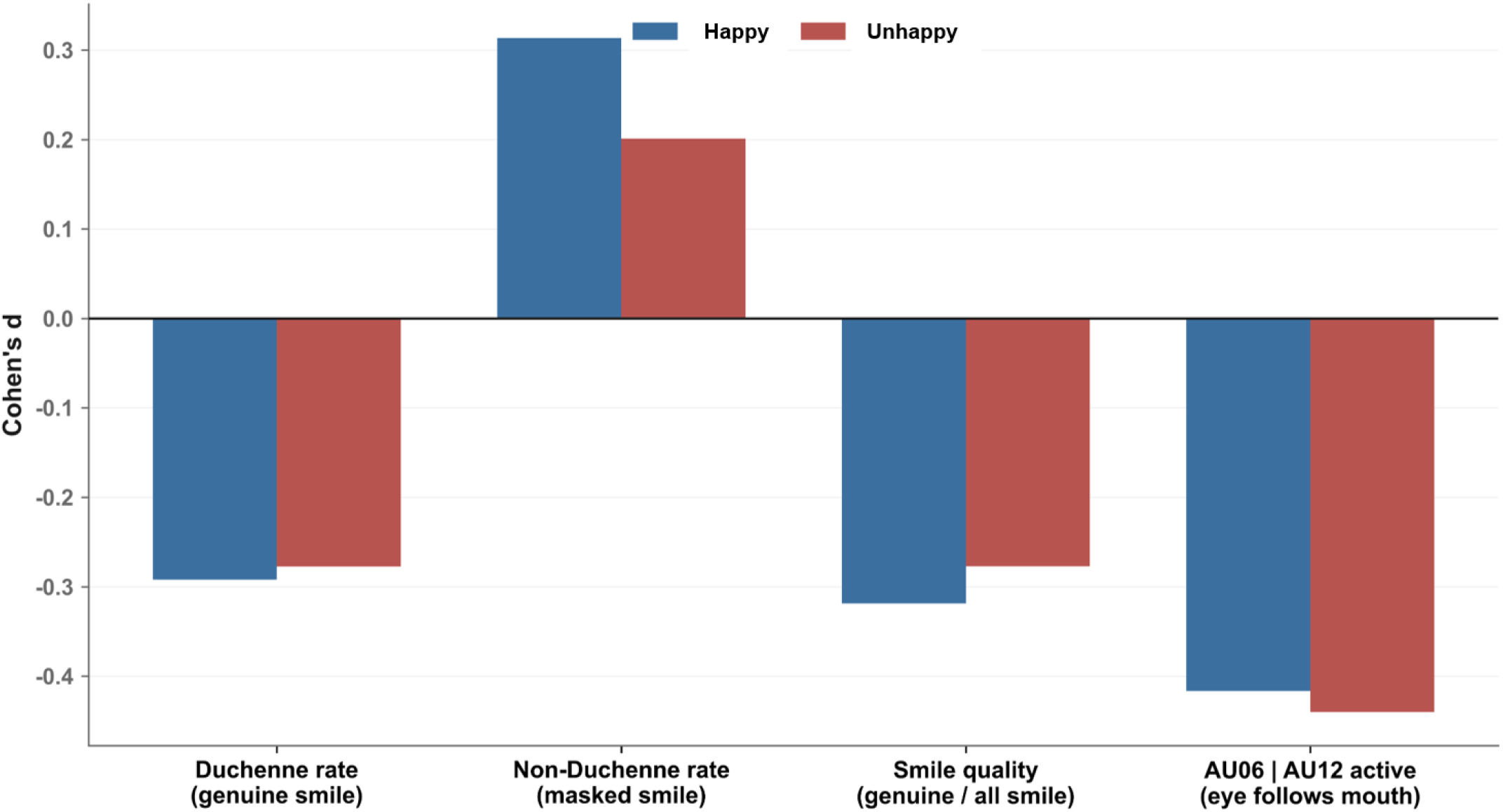
Effect sizes of core smile metrics. Cohen’s d for the four core smile metrics (Duchenne rate, Non-Duchenne rate, smile quality, and au06_given_au12) in the happy and unhappy conditions. Negative values indicate lower values in depressed participants; positive values indicate higher. Asterisks denote FDR-significant group differences. The eye-mouth decoupling index (au06_given_au12) shows the largest effect in both conditions.

**Supplementary Fig. S4.**
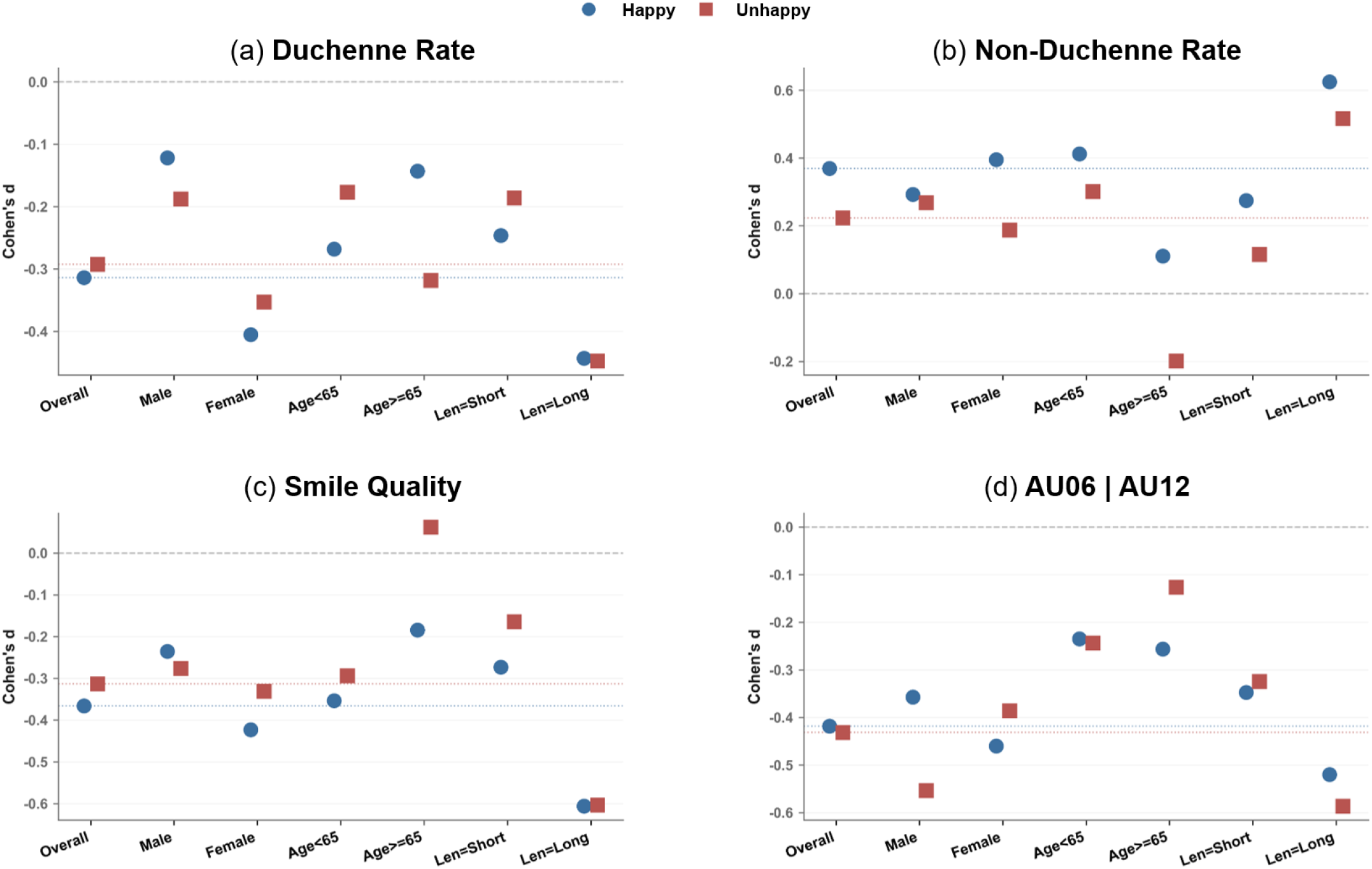
Core smile metrics across subgroups. (a) Duchenne rate, (b) non-Duchenne rate, (c) smile quality, and (d) au06_given_au12 across the overall sample and within sex, age, and speech-length subgroups, shown separately for the happy and unhappy conditions. Cohen’s d values are displayed for each subgroup, and dotted horizontal lines indicate the corresponding overall effect size. The eye–mouth decoupling index (au06_given_au12) remained negative across all subgroups, indicating consistent directionality across sex, age, and speech-length strata. Effect-size estimates from subgroups with small numbers of depressed participants should be interpreted with caution.

**Supplementary Fig. S5.**
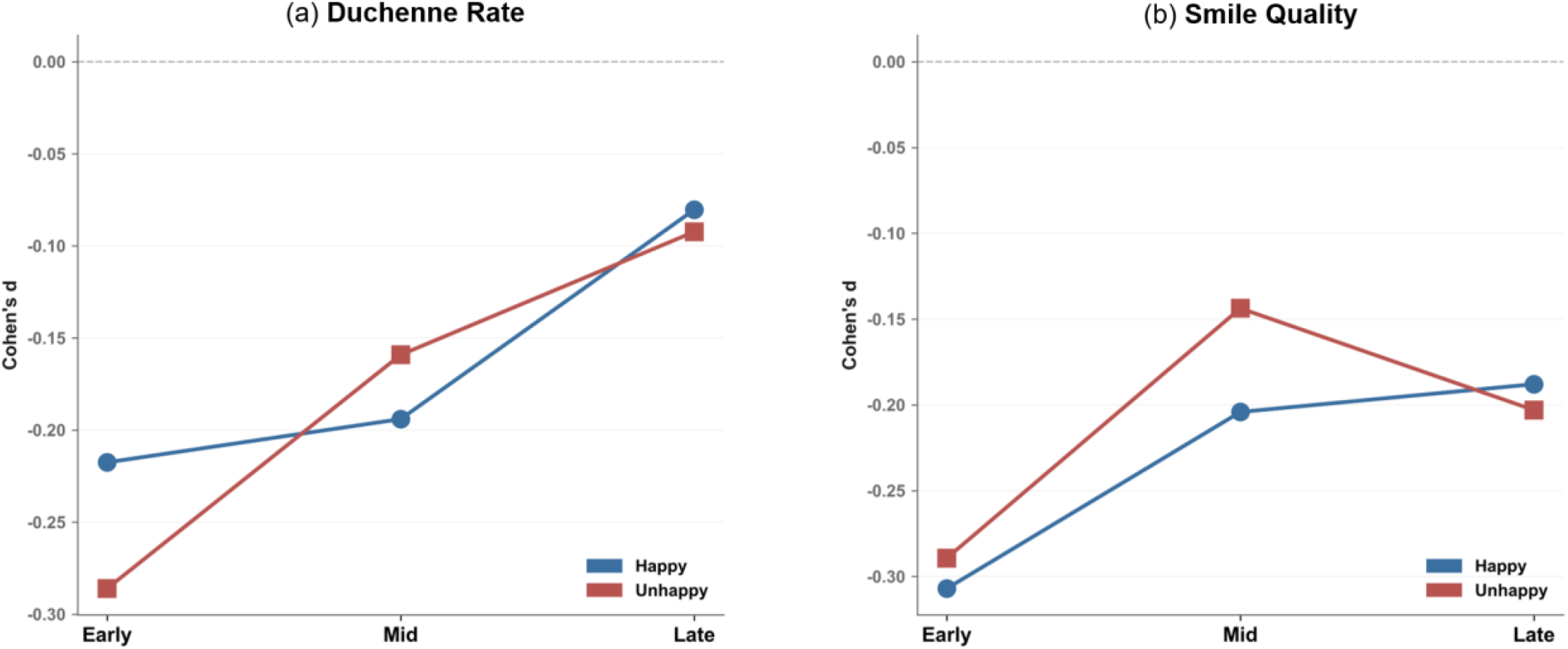
Temporal course of genuine smiling across session thirds, by condition. (a) Duchenne rate and (b) smile quality across relative session thirds (early, mid, late), shown separately for the happy and unhappy conditions. Cohen’s d values are presented for each temporal segment. Group differences were largest in the early segment and generally smaller in later segments, consistent with the temporal patterns described in the main text. Segments represent relative thirds of valid frames rather than absolute elapsed time.

## Notes

This work was supported by the National Research Foundation of Korea (NRF), funded by the Korean Government (MSIT), under Grant RS-2025-00519776 and Grant RS-2024-00440371.

### Competing Interest Statement

The authors have declared no competing interest.

### Author Declarations

This study was based on a single Korean cohort. Participants were recruited at Chonnam National University Hospital (CNUH) and Chonnam National University Hwasun Hospital (CNUHH) from August 2, 2021, to September 26, 2025, as part of a study designed to investigate mental disorders using a transdiagnostic approach based on common symptoms and processes. The study was approved by the CNUH and CNUHH Institutional Review Boards (approval nos. CNUH-2021-243, CNUH-2022-216, CNUHH-2021-117, and CNUHH-2022-126), and written informed consent was obtained from all participants.

### Summary of Updates

Prior to journal submission, a final review was conducted to correct typographical errors. Furthermore, concise interpretations were integrated into several sections to strengthen the justification of specific claims.

## References

[1] World Health Organization. “Depressive disorder (depression),” Accessed: Jun. 30, 2026. [Online]. Available: https://www.who.int/news-room/fact-sheets/detail/depression.

[2] J. Radez, T. Reardon, C. Creswell, F. Orchard, and P. Waite, “Adolescents’ perceived barriers and facilitators to seeking and accessing professional help for anxiety and depressive disorders: a qualitative interview study,” European child & adolescent psychiatry, vol. 31, no. 6, pp. 891–907, 2022.

[3] Z. Liu, B. Hu, L. Yan, T. Wang, F. Liu, X. Li, and H. Kang, “Detection of depression in speech,” in 2015 international conference on affective computing and intelligent interaction (ACII), 2015, pp. 743–747.

[4] S. Byun, A. Y. Kim, E. H. Jang, S. Kim, K. W. Choi, Y. Yu, and H. J. Jeon, “Detection of major depressive disorder from linear and nonlinear heart rate variability features during mental task protocol,” Computers in biology and medicine, vol. 112, pp. 103381, 2019.

[5] M. Jhon, J.-W. Kim, K. Lee, D. Kim, S.-H. Park, C. Kim, B. Lim, S.-Y. Kim, S.-W. Kim, and J.-M. Kim, “Contactless depression screening via facial video-derived heart rate variability,” Translational Psychiatry, 2026.

[6] J. F. Cohn, T. S. Kruez, I. Matthews, Y. Yang, M. H. Nguyen, M. T. Padilla, F. Zhou, and F. De la Torre, “Detecting depression from facial actions and vocal prosody,” in 2009 3rd international conference on affective computing and intelligent interaction and workshops, 2009, pp. 1–7.

[7] J. M. Girard, J. F. Cohn, M. H. Mahoor, S. M. Mavadati, Z. Hammal, and D. P. Rosenwald, “Nonverbal social withdrawal in depression: Evidence from manual and automatic analyses,” Image and vision computing, vol. 32, no. 10, pp. 641–647, 2014.

[8] P. Ekman, E. R. Sorenson, and W. V. Friesen, “Pan-cultural elements in facial displays of emotion,” Science, vol. 164, no. 3875, pp. 86–88, 1969.

[9] P. Ekman, and W. V. Friesen, Facial action coding system: Consulting Psychologists Press, Palo Alto, CA, 1978.

[10] T. Baltrusaitis, A. Zadeh, Y. Lim, and L. Morency, “OpenFace 2.0: Facial Behavior Analysis Toolkit,” in 2018 13th IEEE International Conference on Automatic Face & Gesture Recognition (FG 2018), 2018, pp. 59–66.

[11] Y. Mahayossanunt, N. Nupairoj, S. Hemrungrojn, and P. Vateekul, “Explainable depression detection based on facial expression using LSTM on attentional intermediate feature fusion with label smoothing,” Sensors, vol. 23, no. 23, pp. 9402, 2023.

[12] A. Parikh, M. Sadeghi, R. Richer, L. H. Rupp, L. Schindler-Gmelch, M. Keinert, M. Hager, K. Capito, F. Rahimi, and B. Egger, “Exploring facial biomarkers for depression through temporal analysis of action units,” arXiv preprint arXiv:2407.13753, 2024.

[13] J. Jeganathan, R. Thienel, and M. Breakspear, “Quantifying facial affect changes in psychotic disorders with machine learning,” Psychiatry Research, pp. 116768, 2025.

[14] X. Xu, X. Zhang, and Y. Zhang, “Faces of the Mind: Unveiling Mental Health States Through Facial Expressions in 11,427 Adolescents,” IEEE Transactions on Affective Computing, 2026.

[15] P. Ekman, R. J. Davidson, and W. V. Friesen, “The Duchenne smile: Emotional expression and brain physiology: II,” Journal of personality and social psychology, vol. 58, no. 2, pp. 342, 1990.

[16] M. G. Frank, P. Ekman, and W. V. Friesen, “Behavioral markers and recognizability of the smile of enjoyment,” Journal of personality and social psychology, vol. 64, no. 1, pp. 83, 1993.

[17] G. E. Schwartz, P. L. Fair, P. Salt, M. R. Mandel, and G. L. Klerman, “Facial muscle patterning to affective imagery in depressed and nondepressed subjects,” Science, vol. 192, no. 4238, pp. 489–491, 1976.

[18] J. Gratch, R. Artstein, G. M. Lucas, G. Stratou, S. Scherer, A. Nazarian, R. Wood, J. Boberg, D. DeVault, and S. Marsella, “The distress analysis interview corpus of human and computer interviews,” in Proceedings of the 9th International Conference on Language Resources and Evaluation (LREC), 2014, pp. 3123–3128.

[19] H. Kim, S. Kwak, S. Y. Yoo, E. C. Lee, S. Park, H. Ko, M. Bae, M. Seo, G. Nam, and J.-Y. Lee, “Facial expressions track depressive symptoms in old age,” Sensors, vol. 23, no. 16, pp. 7080, 2023.

[20] K. Mao, Y. Wu, and J. Chen, “A systematic review on automated clinical depression diagnosis,” Npj mental health research, vol. 2, no. 1, pp. 20, 2023.

[21] S. V. Patapati, I. Pendyala, M. Ambati, P. Kunadharaju, P. Kokati, A. Adiraju, and T. Srinivasan, “Most DAIC-WOZ Depression Classifiers Are Invalid, They Don’t Learn Task-Specific Features: Preliminary Findings From a Large-Scale Reproducibility Study,” in Companion Proceedings of the 27th International Conference on Multimodal Interaction, 2025, pp. 17–21.

[22] H. Fisher, P. T. Reiss, D. Atias, M. Malka, B. Shahar, S. Shamay-Tsoory, and S. Zilcha-Mano, “Facing emotions: Between-and within-sessions changes in facial expression during psychological treatment for depression,” Clinical Psychological Science, vol. 12, no. 5, pp. 840–854, 2024.

[23] M. Wang, Y. Wu, K. Rezaee, and M. Khosravi, “Optimized CNN-based facial analysis for depression detection: managing mental disorder in education,” Brain Research Bulletin, pp. 111829, 2026.

[24] R. E. Jack, O. G. Garrod, H. Yu, R. Caldara, and P. G. Schyns, “Facial expressions of emotion are not culturally universal,” Proceedings of the National Academy of Sciences, vol. 109, no. 19, pp. 7241–7244, 2012.

[25] E. Lim, M. Jhon, J.-W. Kim, S.-H. Kim, S. Kim, and H.-J. Yang, “A lightweight approach based on cross-modality for depression detection,” Computers in Biology and Medicine, vol. 186, pp. 109618, 2025.

[26] M. Gonzalez-Machorro, U. Reichel, P. Hecker, H. Hammer, H. Sagha, F. Eyben, R. Hoepner, and B. W. Schuller, “Speech-based depressive mood detection in the presence of multiple sclerosis: A cross-corpus and cross-lingual study,” arXiv, 2025.

[27] Y. Yang, J. Lyu, R. Wang, Q. Wen, L. Zhao, W. Chen, S. Bi, J. Meng, K. Mao, and Y. Xiao, “A digital mask to safeguard patient privacy,” Nature medicine, vol. 28, no. 9, pp. 1883–1892, 2022.

[28] C. Yang, X. Zhang, Y. Chen, Y. Li, S. Yu, B. Zhao, T. Wang, L. Luo, and S. Gao, “Emotion-dependent language featuring depression,” Journal of Behavior Therapy and Experimental Psychiatry, vol. 81, pp. 101883, 2023.

[29] C. Hitchcock, J. Newby, E. Timm, R. M. Howard, A.-M. Golden, W. Kuyken, and T. Dalgleish, “Memory category fluency, memory specificity, and the fading affect bias for positive and negative autobiographical events: Performance on a good day–bad day task in healthy and depressed individuals,” Journal of Experimental Psychology: General, vol. 149, no. 1, pp. 198, 2020.

[30] K. Kroenke, R. L. Spitzer, and J. B. Williams, “The PHQ-9: validity of a brief depression severity measure,” Journal of general internal medicine, vol. 16, no. 9, pp. 606–613, 2001.

[31] X. Li, X. Yi, L. Lu, H. Wang, Y. Zheng, M. Han, and Q. Wang, “TSFFM: Depression detection based on latent association of facial and body expressions,” Computers in Biology and Medicine, vol. 168, pp. 107805, 2024.

[32] M. Sadeghi, R. Richer, B. Egger, L. Schindler-Gmelch, L. H. Rupp, F. Rahimi, M. Berking, and B. M. Eskofier, “Harnessing multimodal approaches for depression detection using large language models and facial expressions,” npj Mental Health Research, vol. 3, no. 1, pp. 66, 2024.

[33] S. Li, and W. Deng, “A deeper look at facial expression dataset bias,” IEEE Transactions on Affective Computing, vol. 13, no. 2, pp. 881–893, 2020.

[34] J. F. Cohn, I. O. Ertugrul, W.-S. Chu, J. M. Girard, L. A. Jeni, and Z. Hammal, “Affective facial computing: Generalizability across domains,” Multimodal Behavior Analysis in the Wild, pp. 407–441: Elsevier, 2019.

[35] I. O. Ertugrul, J. F. Cohn, L. A. Jeni, Z. Zhang, L. Yin, and Q. Ji, “Crossing domains for au coding: Perspectives, approaches, and measures,” IEEE transactions on biometrics, behavior, and identity science, vol. 2, no. 2, pp. 158–171, 2020.

[36] T. Chen, T. Pu, H. Wu, Y. Xie, L. Liu, and L. Lin, “Cross-domain facial expression recognition: A unified evaluation benchmark and adversarial graph learning,” IEEE transactions on pattern analysis and machine intelligence, vol. 44, no. 12, pp. 9887–9903, 2021.

[37] J. Gideon, M. G. McInnis, and E. M. Provost, “Improving cross-corpus speech emotion recognition with adversarial discriminative domain generalization (ADDoG),” IEEE Transactions on Affective Computing, vol. 12, no. 4, pp. 1055–1068, 2019.

[38] R. Wang, J. Huang, J. Zhang, X. Liu, X. Zhang, Z. Liu, P. Zhao, S. Chen, and X. Sun, “Facialpulse: An efficient rnn-based depression detection via temporal facial landmarks,” in Proceedings of the 32nd ACM international conference on multimedia, 2024, pp. 311–320.

[39] J. M. Girard, G. Shandar, Z. Liu, J. F. Cohn, L. Yin, and L.-P. Morency, “Reconsidering the duchenne smile: indicator of positive emotion or artifact of smile intensity?,” in 2019 8th International Conference on Affective Computing and Intelligent Interaction (ACII), 2019, pp. 594–599.

[40] R. M. Müri, “Cortical control of facial expression,” Journal of comparative neurology, vol. 524, no. 8, pp. 1578–1585, 2016.

[41] W. E. Rinn, “The neuropsychology of facial expression: a review of the neurological and psychological mechanisms for producing facial expressions,” Psychological bulletin, vol. 95, no. 1, pp. 52, 1984.

[42] K. M. Gothard, “The amygdalo-motor pathways and the control of facial expressions,” Frontiers in neuroscience, vol. 8, pp. 43, 2014.

[43] L. M. Bylsma, B. H. Morris, and J. Rottenberg, “A meta-analysis of emotional reactivity in major depressive disorder,” Clinical psychology review, vol. 28, no. 4, pp. 676–691, 2008.

[44] P. H. Mui, M. B. Goudbeek, M. G. Swerts, and A. Hovasapian, “Children’s nonverbal displays of winning and losing: Effects of social and cultural contexts on smiles,” Journal of Nonverbal Behavior, vol. 41, no. 1, pp. 67–82, 2017.

[45] F. Ringeval, B. Schuller, M. Valstar, N. Cummins, R. Cowie, L. Tavabi, M. Schmitt, S. Alisamir, S. Amiriparian, and E.-M. Messner, “AVEC 2019 workshop and challenge: state-of-mind, detecting depression with AI, and cross-cultural affect recognition.” pp. 3–12, 2019.

[46] N. Jin, R. Ye, and P. Li, “Diagnosis of depression based on facial multimodal data,” Frontiers in psychiatry, vol. 16, pp. 1508772, 2025.

[47] N. Cummins, S. Scherer, J. Krajewski, S. Schnieder, J. Epps, and T. F. Quatieri, “A review of depression and suicide risk assessment using speech analysis,” Speech communication, vol. 71, pp. 10–49, 2015.

[48] H. Fisher, N. M. Jaffe, K. Pidvirny, A. O. Tierney, M. S. Vaidean, P. Dongre, and C. A. Webb, “Language-based detection of depression with machine learning: systematic review and meta-analysis,” npj Digital Medicine, 2026.

